# Identifying gaps in tuberculosis preventive care for non-U.S.-born persons at community health clinics in the United States

**DOI:** 10.1101/2025.06.24.25330215

**Authors:** Priya B. Shete, Matthew T. Murrill, Katharine M. Tatum, Amina Ahmed, Kaylynn Aiona, Jeannette L. Aldous, Jennifer Flood, Michelle K. Haas, Masahiro Narita, April C. Pettit, Marie N. Séraphin, Adithya Cattamanchi, Tracy L. Ayers, the Tuberculosis Epidemiologic Studies Consortium

## Abstract

**Background:** Treating asymptomatic tuberculosis (TB) infection prevents TB disease and is critical for TB elimination in the United States (U.S.). In the U.S., TB disease disproportionately affects those born outside of the U.S. The quality of TB preventive care in primary care settings for non-U.S.-born populations is poorly characterized. We aimed to measure gaps in TB preventive care among non-U.S.-born persons receiving primary care at community health clinics across the U.S.

**Methods:** We conducted a retrospective cohort study at 12 community health clinics in the U.S. Per clinic, individual-level demographic and TB clinical data were extracted for approximately 700 non-U.S.-born persons aged ≥18 years seeking care between June and December 2019. We constructed a care cascade describing gaps in TB infection testing and treatment and used multivariable mixed effects logistic models to evaluate associations between individual and clinic-level characteristics and observed gaps.

**Results:** Among 8,460 non-U.S.-born individuals included, 68% were female, and the median age was 50 years (interquartile range 38-63). Of those included, 2,765 (33%) had a TB infection test ordered. Among 1,022 with a positive test, 787 (77%) were diagnosed with TB infection, of whom 377 (48%) were offered preventive treatment. Among 173 who were treated at study clinics, 141 (82%) completed treatment.

**Conclusions:** Gaps in testing and treatment initiation for non-U.S.-born individuals were pervasive at the 12 community health clinics in this study. Research is needed to identify strategies that increase TB preventive care among populations who face the disproportionate burden of disease.

**Article summary:** In this multi-site study of U.S. community health clinics, substantial gaps in tuberculosis infection testing and treatment initiation were identified for non-U.S.-born persons, a population that bears the disproportionate burden of TB disease in the U.S.

## Introduction

Treating asymptomatic Mycobacterium tuberculosis (TB) infection (latent tuberculosis infection, LTBI) is critical to preventing TB disease in the United States (U.S.).^1^ After decades of progress, TB disease incidence has increased annually since the COVID-19 pandemic, highlighting the urgency of TB prevention.^2^ An estimated 13 million people in the U.S. (5% of the population in 2011-2012) may have TB infection with more than 80% of TB disease diagnosed in the U.S. due to the reactivation of LTBI.^3^ Modeling studies indicate that substantial increases in LTBI testing and treatment are needed to approach U.S. TB elimination goals (<1 TB disease diagnosis/million persons/year) by 2050.^4,5^

Over 70% of people diagnosed with TB disease in the U.S. were born or lived outside of the U.S. in high TB incidence settings,^2^ with over 90% of these diagnoses attributed to the reactivation of LTBI acquired prior to immigration.^6^ Non-U.S.-born individuals in the U.S. are more than 10 times likely to have LTBI compared to U.S.-born individuals.^2^ Notably, half of TB disease among non-U.S.-born persons is diagnosed 10+ years after U.S. arrival.^7^ Given these data, multiple guidelines recommend testing all non-U.S.-born persons from high-TB incidence settings for TB infection, regardless of time since U.S. arrival.^8,9^

TB elimination in the U.S. requires the scale-up of TB infection testing and treatment beyond public health clinics, which alone lack the capacity and resources,^10^ to primary care clinics. The expansion of insurance coverage of preventive care services through the Affordable Care Act,^11^ the availability of more specific interferon-gamma release assay (IGRA) tests [i.e., QuantiFERON (QFT) and T-SPOT.TB (T-SPOT)] and safer rifampin-containing TB preventive treatment regimens^12^ all have the potential to accelerate TB elimination efforts through primary care. While most individuals receive routine medical care in outpatient primary care clinics, these clinics, however, may not all be currently equipped to provide TB and LTBI care.^13,14^

The diagnosis and treatment of LTBI can be characterized as a cascade of care starting with individuals at increased risk (e.g., non-U.S.-born persons) eligible for screening^9^ to treatment completion. Systematic reviews have highlighted large gaps in TB infection testing and treatment for persons migrating from high to low-TB incidence settings with a wide range of barriers to guideline-directed care.^15,16^ However, there are limited data on the gaps in care for non-U.S.-born persons from geographically diverse primary care clinics in the U.S.^17,18^

The objectives of this study are to describe the LTBI care cascade among non-U.S.-born individuals in primary care and identify health systems and individual-level barriers and facilitators to high-quality LTBI care.

## Methods

### Study setting

We conducted a retrospective cohort study at 12 community health clinics (CHCs) (**Figure E1 in Supplementary Materials**) participating in the Tuberculosis Epidemiologic Studies Consortium 2 (TBESC-2) across 10 states in the U.S.^19^ CHCs were defined as any general or family medicine primary care clinic not run by a state or local health department that served an adult population. CHCs were eligible to participate if they: 1) were involved in TBESC-2, 2) provided routine care to ≥25 individuals daily from intermediate- to high-TB incidence countries (i.e., all countries except United States, Canada, Australia, Japan, New Zealand, and countries of Western Europe) and 3) provided LTBI diagnostic services.

### Study population

All individuals presenting for routine care at each clinic on 4 randomly selected days per month between June 2019 to December 2019 were screened for inclusion into the cohort. Individuals were included based on the below eligibility criteria until a goal of 700 individuals per clinic was reached. Inclusion criteria were: 1) adult (≥18 years of age), 2) presentation for care on days of data collection, 3) established care at the clinic for ≥12 months and 4) non-U.S.-born, defined as birth outside of the U.S. or non-English primary language or use of an interpreter during a medical visit if place of birth was not available. Non-English primary language has been shown to have high specificity but limited sensitivity for birth outside of the U.S.^20,21^

### Data collection

After identification of eligible individuals, TBESC-2 staff visited each CHC to review paper or electronic medical records to confirm eligibility and extract demographic and clinical data using a standardized form (**Supplementary Materials**). Chart review started with the individual’s most recent visit and proceeded retrospectively to include their entire history at the clinic. Specifically, data on TB infection testing and treatment were extracted, including reasons treatment was not prescribed, started or completed. Information on clinic characteristics and TB infection testing and treatment practices was collected through interviews with clinic staff using a standardized assessment (**Supplementary Materials**).

### Construction of LTBI care cascade

Definitions for each step in the LTBI care cascade were based on recent systematic reviews^15,16^ and included a total of eight steps, starting with eligibility for testing and ending in LTBI treatment completion (**Table 1**). Definitions for each cascade step are included in **Supplementary Materials**.

**Table 1:**
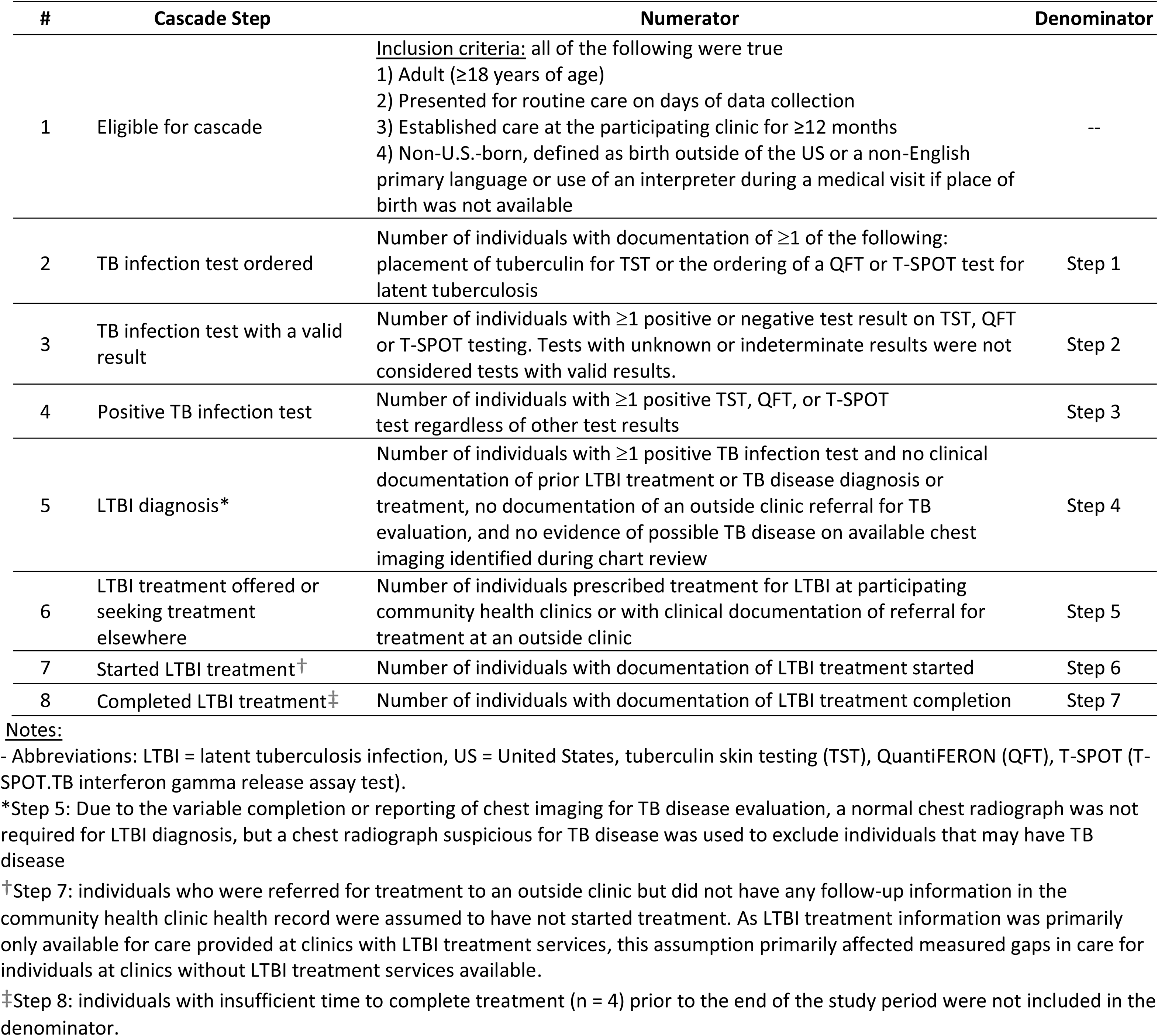
Definitions for steps in the latent tuberculosis infection care cascade, including numerator and denominator values used to calculate the proportion of individuals successfully completing each step.

### Statistical analysis

For the construction of the LTBI care cascade, two indicators for each step were reported: number of individuals meeting criteria for each step and the percentage of individuals from a prior step that progressed to the subsequent step. To evaluate associations of individual-level and clinic-level factors with the completion of key cascade steps (ordering TB infection test, positive TB infection test, prescribed LTBI treatment or seeking treatment elsewhere, completing LTBI treatment), multivariable mixed effects logistic regression models with a random intercept for clinic were fit to account for clinic-level clustering of data. Informed by literature review, prior qualitative interviews, and available data, individual and clinic-level covariates were selected for each cascade step model prior to analysis (**Supplementary Materials**). Empirical Bayes estimates of the clinic-level random effects for each outcome were plotted to assess clinic-level variability. Stratified LTBI care cascades were also constructed by patterns of tests used to evaluate for LTBI [e.g. tuberculin skin test (TST), IGRA, or combination) and their results. All statistical analyses were conducted in Stata v17 (StataCorp, College Station, TX, USA).

### Human research ethics approvals

This study was reviewed and approved by the CDC Institutional Review Board (IRB) and University of California, San Francisco IRB (IRB#18-27014).

## Results

### Study population

From 12 CHCs, 8,460 non-U.S.-born individuals were included in the analysis with a median of 702 individuals per site (range 696-735) (**Table 2**). Four people were excluded due to missing age (n=2) and LTBI care cascade eligibility criteria (n=2). Among those included, the median age was 50 years [interquartile range (IQR) 38-63 years, range 18-97 years], and 68% were female with clinic-level variability for both demographic variables (data not shown). The top 3 primary languages were Spanish (43%), English (21%), and Cantonese (12%). Of note, there was considerable missing data in the medical record on place of birth (58%), race (38%) and ethnicity (20%). Of the 12 CHCs in the study, 9 (75%) used IGRA for testing and 7 (58%) provided both LTBI diagnostic and treatment services (**Table E1**).

**Table 2:**
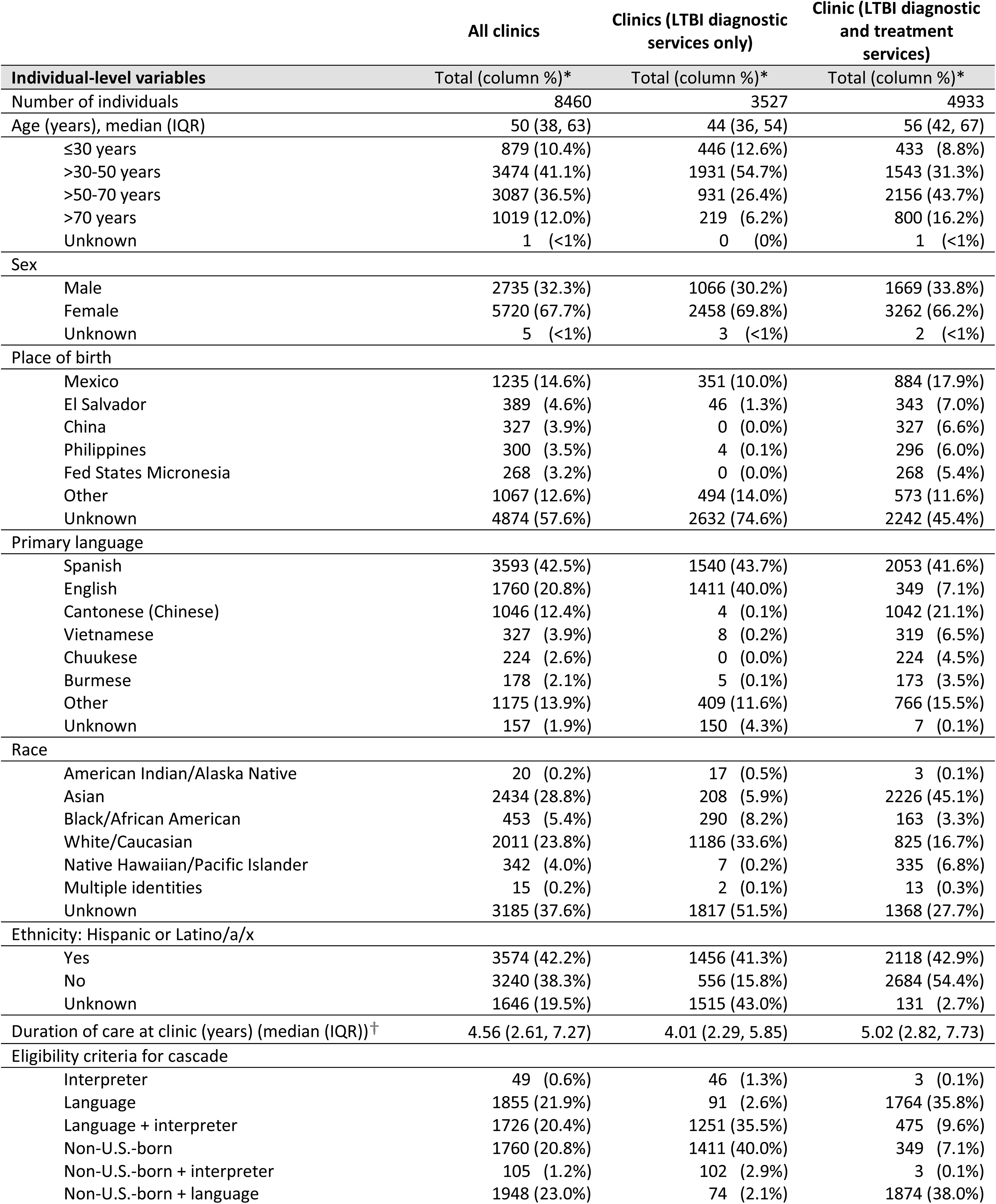
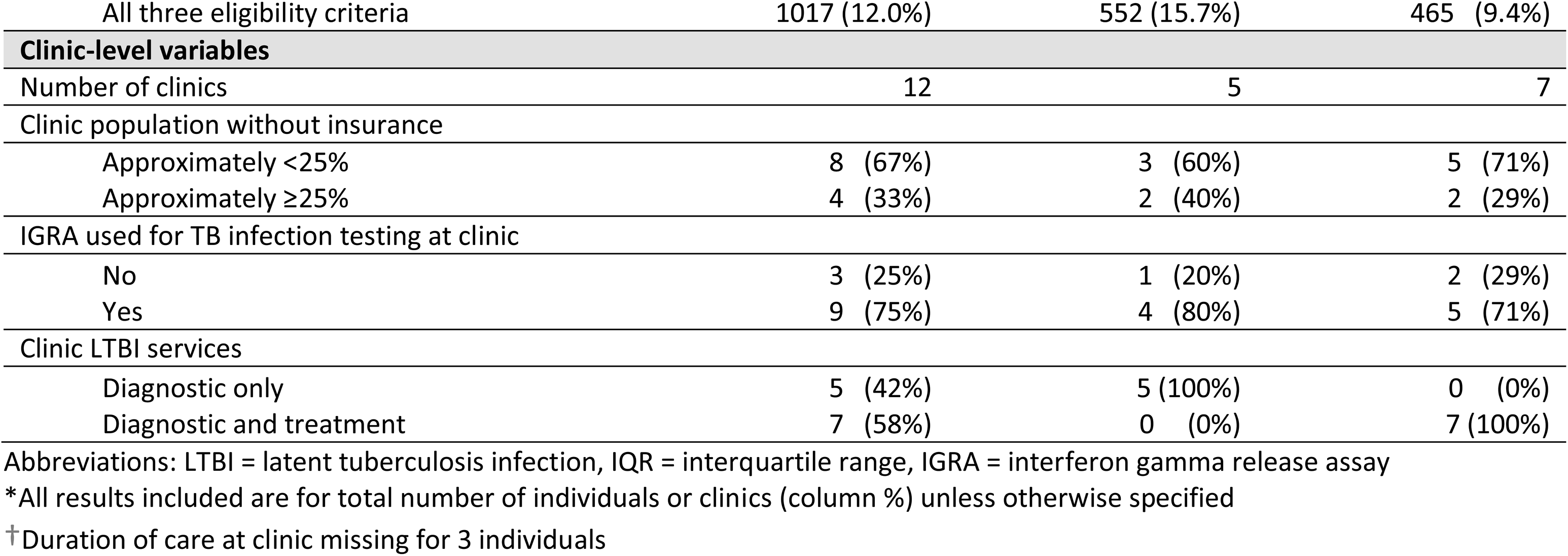
Clinical and demographic characteristics of non-U.S.-born individuals by community health clinics with varying latent TB infection services provided. Characteristics of participating community health clinics.

### Overall LTBI care cascade

The LTBI care cascade with reasons for losses between each step for all 12 CHCs is presented in **Figure 1** and stratified by the availability of LTBI clinic services in **Figure E2**. For all clinics, the LTBI diagnostic cascade stratified by clinic is presented in **Figure E3.** For clinics offering LTBI treatment, the entire care cascade stratified by clinic is presented in **Figure E4**.

**Figure 1:**
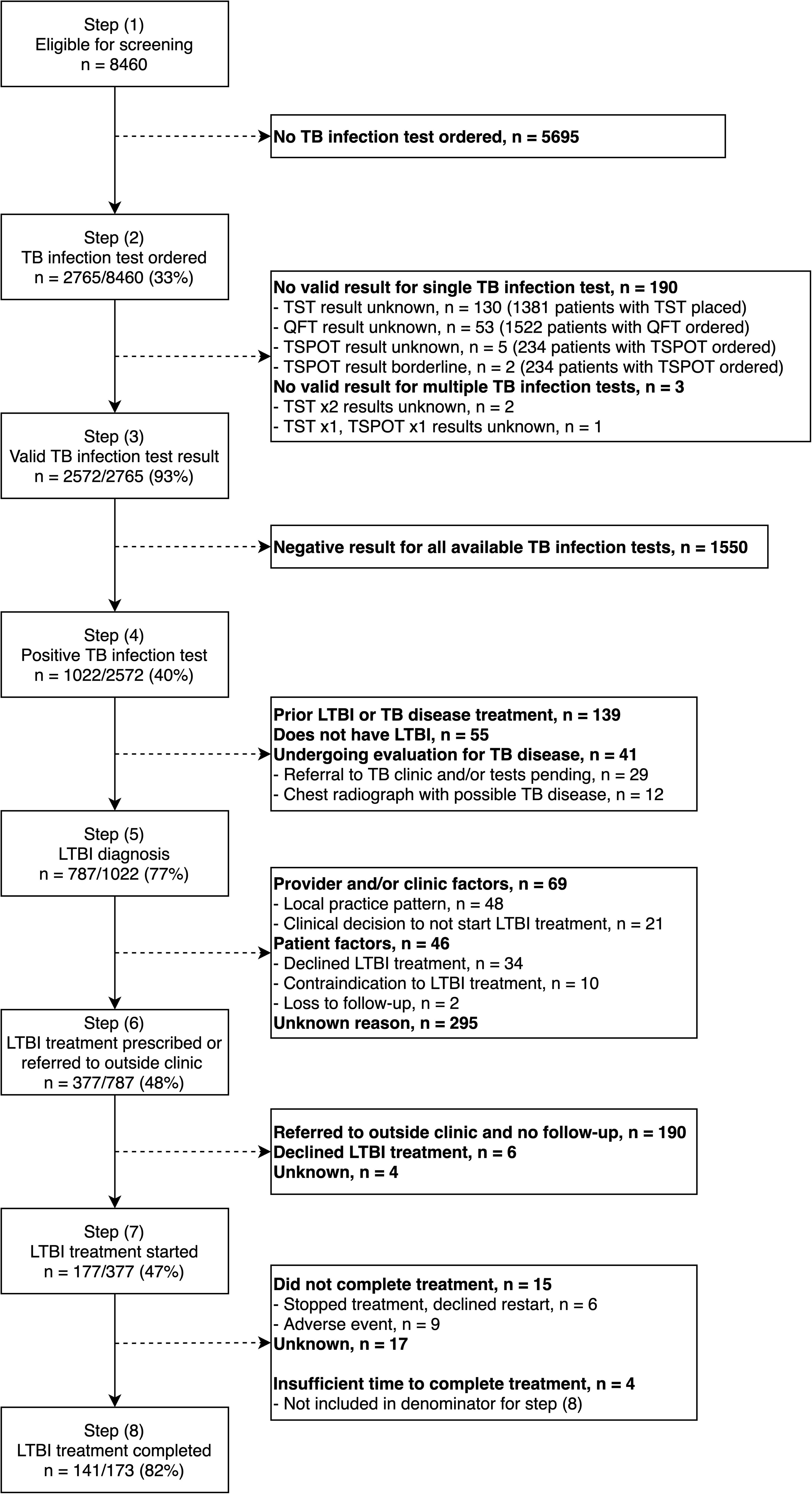
Eligibility and latent TB infection care cascade for non-U.S.-born individuals at 12 participating community health clinics across the United States. Abbreviations: n = number of individuals, TB = tuberculosis, LTBI = latent tuberculosis infection, tuberculin skin testing (TST), QuantiFERON (QFT), T-SPOT (T-SPOT.TB interferon gamma release assay test)

### TB infection testing (cascade steps 2-4)

Among 8,460 non-U.S.-born individuals included in the study, 2,765 (33%) had at least one TB infection test ordered (TST or IGRA). Multiple tests were ordered for 447 individuals (16% of 2,765). Of the 2,765 individuals, 48% had TST placed, 55% had QFT ordered and 3% had T-SPOT ordered. Test use also varied by site (**Table E2**), but data were not systematically available on reasons for individuals not being tested. Factors associated with having at least one TB infection test ordered included: longer duration of care at a clinic [adjusted odds ratio (aOR) 1.07 per one year increase, 95% confidence interval (95%CI) 1.05-1.09] and receiving care at a clinic that provided both LTBI diagnostic and treatment services (aOR 8.87, 95%CI 2.74-28.74). Age older than 30 years was associated with lower odds of a documented TB infection test ordered compared to individuals ≤30 years (**Table 3**).

**Table 3:**
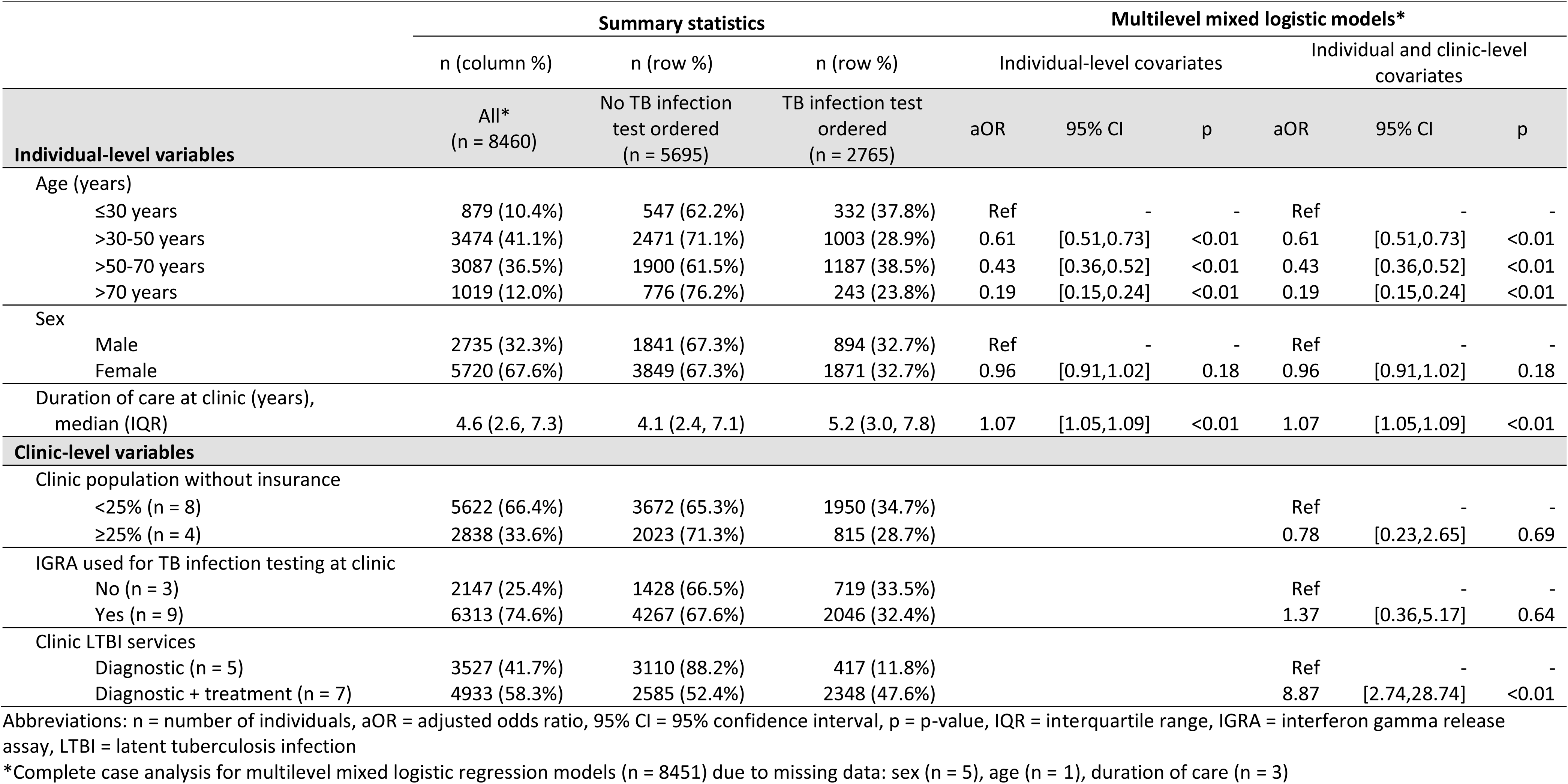
Individual and clinic-level factors associated with TB infection test ordering for non-U.S.-born individuals at 12 community health clinics in the United States.

Among 2,765 individuals who had at least one TST placed or IGRA ordered, 2,572 (93%) had valid results for at least one test, and 444 (16%) had more than one valid test result. Individuals with only a single TST placed were less likely to have a valid test result available (839/969, 87%) compared to those with a single IGRA test ordered (1,289/1,349, 96%) or multiple tests ordered or placed (444/447, 99%).

Of 2,572 individuals with at least one valid test for TB infection, 1,022 (40%) had at least one positive result. Factors associated with increased odds of at least one positive test, included (**Table E3**): age older than 30 years, longer duration of care at a clinic (aOR 1.06 per 1 year increase, 95%CI 1.03-1.09), and more than one test ordered or placed (aOR 4.25, 95%CI 3.34-5.40). Females had lower odds of a positive test than males (aOR 0.74, 95%CI 0.62-0.89) but not of having a test ordered (**Table 3**). There was significant variation in TB infection testing steps across CHCs (**Figures E3 and E5)**: tests ordered if eligible (median clinic 26%, IQR 13-47%), tests completed if ordered (median clinic 95%, IQR 82-99%), and at least one test positive (median clinic 40%, IQR 29-48%).

### LTBI evaluation and diagnosis (cascade step 5)

There was also variability of on-site radiology services at clinics (**Table E4)**, and chest radiographs with results were only available for 705 of the 1,022 (69%) individuals with a positive test for TB infection. Among individuals with a positive test, 787 (77%) were considered to have an LTBI diagnosis after review of any available chest imaging results and clinical notes. On chart review, 139 out of 1,022 individuals (14%) with a positive test had prior TB or LTBI treatment, 55 (5%) did not have LTBI but with unclear final diagnosis, and 41 (4%) were undergoing evaluation for TB disease.

### LTBI treatment (cascade steps 6-8)

As only 7 of 12 CHCs provided on-site LTBI treatment services, the first LTBI treatment cascade step was defined as either treatment prescribed or documentation that an individual was seeking treatment at an outside clinic. Among individuals with an LTBI diagnosis, 48% (377/787) completed this step (**Figure 1**). Provider reasons for not recommending or individual reasons for declining treatment were documented for only 28% (115/410) of persons not prescribed treatment or seeking treatment elsewhere. Approximately 50% (190/377) of individuals prescribed treatment on-site or seeking LTBI treatment elsewhere were referred to an outside clinic with no follow-up information documented. As it was unclear if LTBI treatment was ever started, these individuals were excluded from subsequent cascade steps. Of the remaining individuals, 177 (47% of 377) had documentation of LTBI treatment initiation in their medical record, with whom 172 (97% of 177) received care at CHCs that provided LTBI treatment services on-site. Among individuals who were known to start LTBI treatment, 141 (82% of 173) completed treatment, with 4 individuals removed from the denominator due to insufficient time to complete treatment at the time of chart review. Similar to other cascade steps, significant clinic-level variation was noted for treatment offered (**Figure E6**); however, not for treatment completion (**Figure E7**).

The ordering of IGRA for testing at the individual-level was associated with higher odds of prescription of LTBI treatment or seeking treatment elsewhere (aOR 2.16, 95%CI 1.29-3.64) (**Table 4**). Longer duration of care at a CHC was associated with lower odds of treatment prescribed or seeking treatment elsewhere (aOR 0.93 per 1 year increase, 95%CI 0.88-0.99). In descriptive analyses of 194 individuals with more than one valid TB infection test result and an LTBI diagnosis, those with a positive TST and negative IGRA result were less likely to be offered treatment or referred elsewhere (4/54, 7%) compared to other test result patterns (96/140, 69%) (**Table E5**). In a limited model of individuals on LTBI treatment at 7 participating CHCs where LTBI treatment was prescribed, no individual or clinic-level factors were significantly associated with treatment completion; however, individuals who took 4 months of rifampin (aOR 2.47, 95%CI 0.79-7.67) or other/unknown regimens (aOR 3.34, 95%CI 0.60-18.51) had non-significant but increased odds of treatment completion compared to individuals on 9 months of isoniazid (**Table 5**).

**Table 4:**
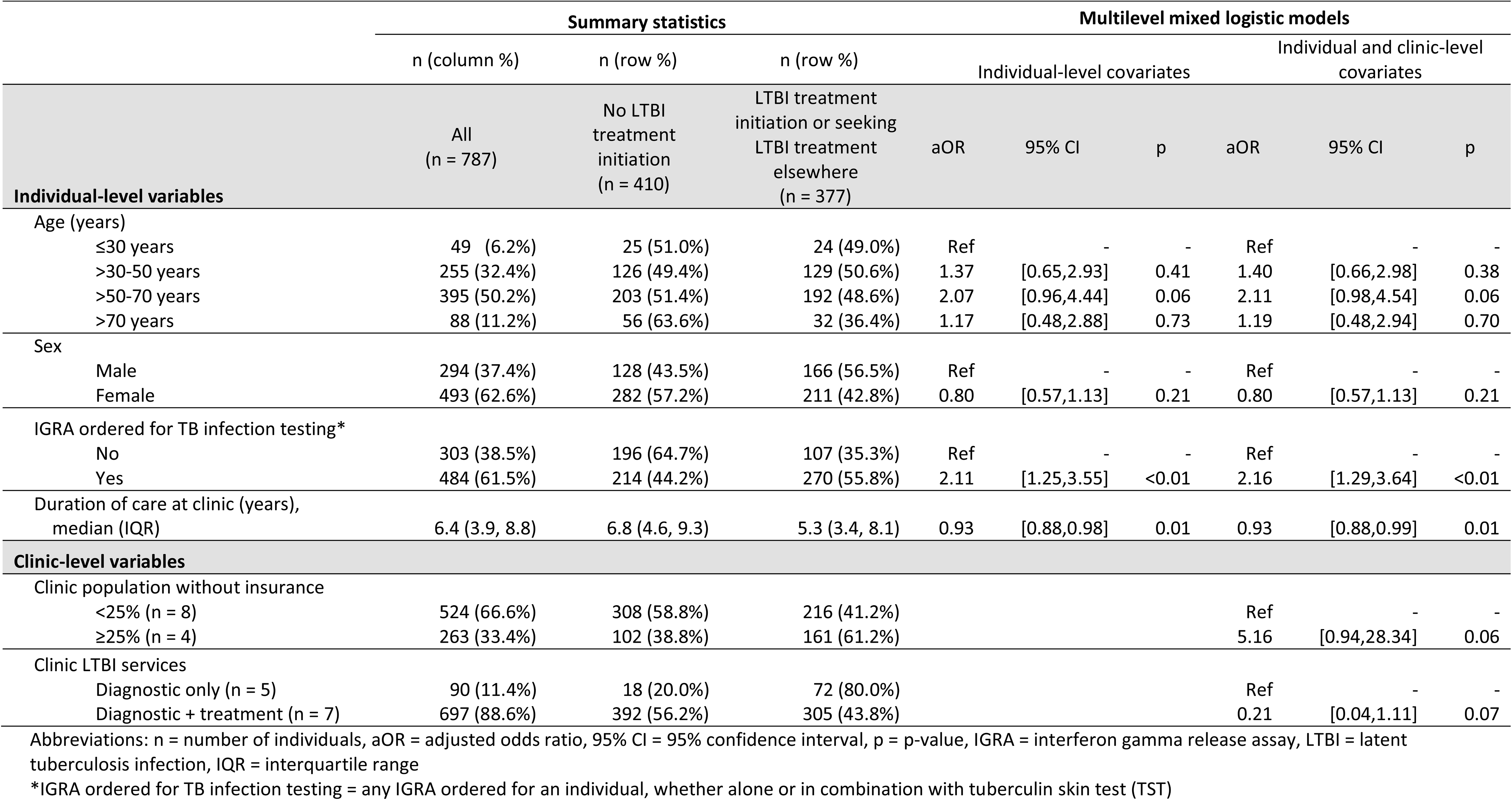
Individual and clinic-level factors associated with LTBI treatment initiation or seeking LTBI treatment elsewhere for non-U.S.-born individuals at 12 community health clinics in the United States.

**Table 5:**
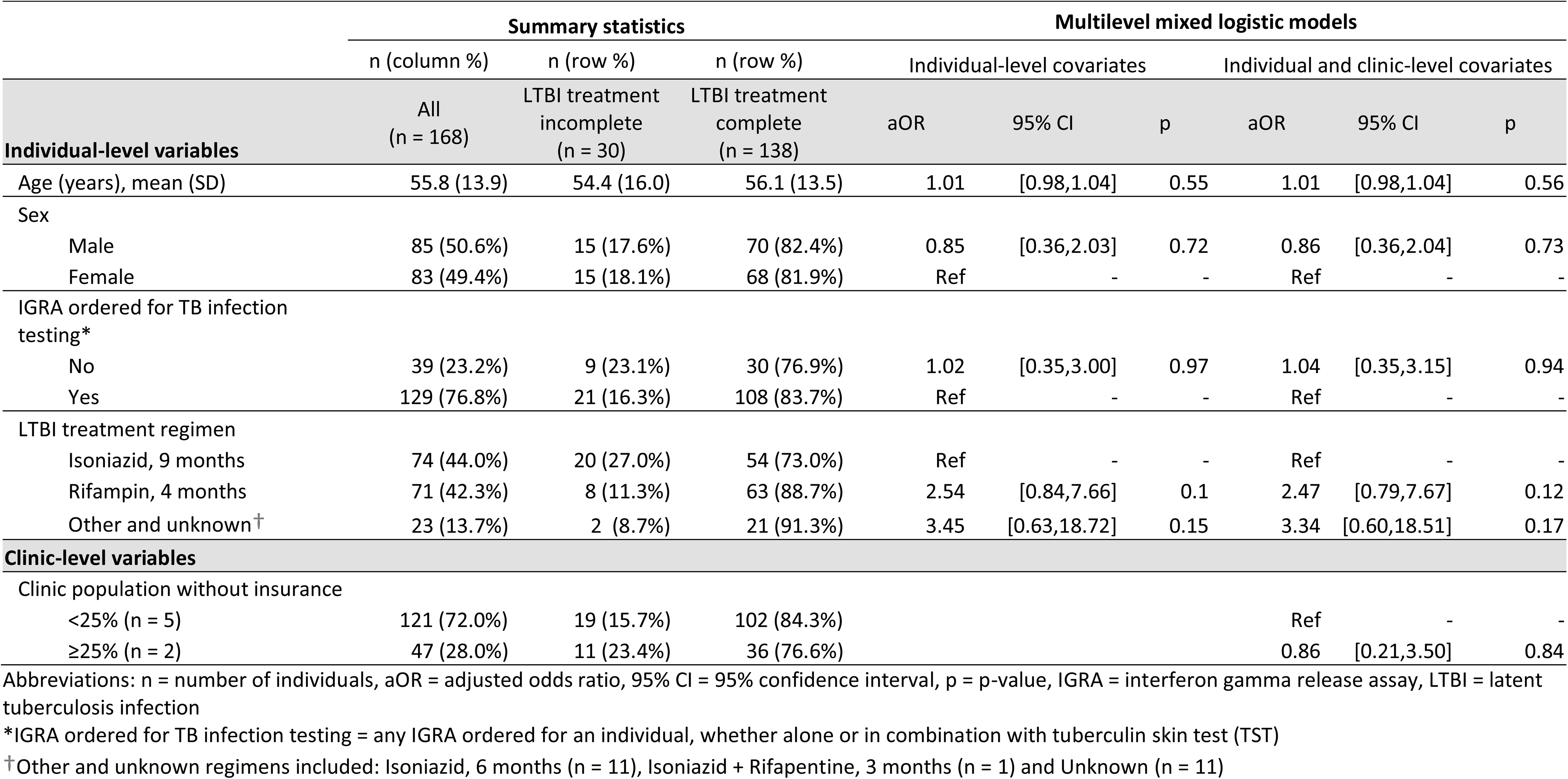
Individual and clinic-level factors associated with LTBI treatment completion for non-U.S.-born individuals at 7 community health clinics in the United States where clinical providers prescribe LTBI treatment.

## Discussion

Globally, tuberculosis is the leading cause of preventable death from an infection. In this multi-site study of CHCs, only one in three non-U.S.-born persons were tested for *M. tuberculosis* infection, despite clinical recommendations for testing since the 1990s.^9^ These findings are similar to LTBI care cascade analyses from a small number of U.S. health systems that identified 13-20% of eligible non-U.S.-born persons were tested,^22,23^ and 37-59% with an LTBI diagnosis were prescribed treatment.^22–24^ An analysis of nationally representative data from the National Health and Nutrition Examination Survey (NHANES) 2011-12 similarly estimated that 22% of individuals with LTBI in the U.S. were diagnosed and 10% have completed treatment.^18^ These findings underscore the need to scale-up TB preventive care.

Although adults ≥65 years of age account for 25% of TB disease in the U.S.,^2^ older adults in the present study had lower odds of testing compared to younger individuals, possibly due to increased medical complexity, competing demands during appointments, concerns about treatment side effects and structural barriers.^25,26^ CHCs offering LTBI diagnostic and treatment services had higher odds of testing compared to those only offering diagnostic services. This association may be due to increased provider knowledge of LTBI management or clinic-level support. While clinic-level resources, such as on-site radiology or pharmacy, may impact the completion of LTBI cascade steps, these were not assessed due to clinic sample size and the overlap in their distribution with clinics offering treatment services.

The prevalence of TB infection test positivity among those tested in this cohort was 40%, which was higher than prior studies from primary care networks in Seattle (29%)^23^ and San Francisco (27%)^22^ as well as large cohorts of public health clinics (20%)^27^ and clinical sites serving people with increased risk of TB (e.g., tuberculosis clinics, student health clinics and refugee programs) (31%).^28^ Analysis of NHANES, 2011-12 data also found a lower prevalence of positive TB infection tests among non-U.S.-born persons (19%).^29^ Possible explanations for this higher test positivity include: 1) individual-level selection bias where LTBI screening is based not just on place of birth but additional risk factors for TB, 2) clinic-level selection bias where participating clinics were associated with other TB research, 3) TB tests ordered for individuals with a prior history of TB disease (139 persons, who if excluded at the cascade eligibility step, would reduce test positivity prevalence to 36% for tested individuals in the cohort), 4) false positive TST results due to Bacillus Calmette-Guérin vaccination, or 5) providers more likely to document prior or outside positive tests over negative ones. Despite these potential biases, it is likely that the true overall prevalence of LTBI among non-U.S.-born persons at CHCs in the present study is at least as high as estimates from prior studies, highlighting the importance of working with CHCs to improve and scale up TB prevention efforts.

Defining LTBI diagnoses using electronic health records is challenging and can vary across cohorts based on data availability.^22–24,27^ In this study, data was not consistently available on TB disease evaluation (e.g., chest imaging and microbiologic tests) or formal LTBI diagnosis (e.g., diagnostic codes). As a result, LTBI diagnosis was defined using a prior positive TB infection test and the absence of clinical documentation of TB disease, an outside referral for TB evaluation, or chest imaging suspicious for TB disease. While the prevalence of TB disease is substantially lower than LTBI in the U.S., it is possible that some individuals with TB disease were misclassified as LTBI.

Among those diagnosed with LTBI, 50% were prescribed treatment or documented as seeking treatment elsewhere. This is consistent with prior U.S. studies of LTBI treatment initiation among non-U.S.-born persons, which ranged between 37-59%.^22–24, 27^ It is also similar to a systematic review of 12 LTBI care cascades in North America and Europe that noted 55% treatment initiation for those with LTBI.^15^ Analyses of treatment completion for this study were limited to the seven provider LTBI treatment services, and treatment outcomes were ascertained through chart review of clinical notes, not medication refill data. Treatment completion (82%) was higher than a recently published cohort from Kaiser Permanente in California (65% completion regardless of place of birth)^24^ and similar to a cohort of local health departments across the U.S. (78% for non-U.S.-born persons).^27^ Uncertainty in these estimates may exist due to missing data, different definitions of treatment completion, or loss to follow-up.

This study had several key strengths. Despite published LTBI care cascades from specific U.S. health systems,^22–24^ the present cohort includes geographically diverse CHCs across the U.S. Ascertainment of treatment outcomes through rigorous chart review was a strength of this work as medication refill data can be limited by missingness or require complex analyses. In addition to the limitations mentioned above, data on demographic variables, such as place of birth, were not available. Similar to all cascade analyses, identified gaps in LTBI care are likely a combination of true gaps, care provided outside of the health system, and a lack of provider documentation.

Overall, this research identified major gaps in LTBI testing and treatment for non-U.S.-born persons at CHCs across the U.S. Ongoing efforts are focused on designing and evaluating interventions to improve the care cascade.^22,30^ However, interventions focusing on single steps of the cascade may “shift attrition downstream”^31^ due to weak linkages, additional barriers to care, and operational challenges.^32^ Integrated and person-centered approaches to addressing access, implementation and quality barriers along the entire cascade have the greatest potential to improve outcomes.^33–35^ Additional research to improve the identification of individuals at highest risk of progression from LTBI to TB has the potential to maximize the benefits of LTBI treatment.^36,37^ There are also ongoing studies of shorter LTBI regimens that may improve treatment completion.^38^

## Supporting information

Clinic Assessment Form

Patient Data Extraction Form

## Disclaimer

The findings and conclusions in this report are those of the authors and do not necessarily represent the official position of the CDC and other funding agencies.

## Acknowledgements

We would like to thank the staff, providers and all individuals receiving care at clinics participating in the present study. We would also like to thank Courtney Coleman, Leah Jarlsberg, Evette Larry, Vanessa Li, Angie Miner, Sarita Mohanty and Matthew Whipple as well as site investigators and site-level research team members for their significant support throughout this project. The authors acknowledge the use of ChatGPT-4o for editorial assistance in suggesting alternative wording in the introduction and discussion sections to reduce the overall word count of the manuscript. The authors manually incorporated possible suggestions from ChatGPT-4o that were made to the text and did not alter the intended meaning.

## CRediT author statements

Priya B. Shete: Conceptualization, Funding acquisition, Investigation, Methodology, Supervision, Writing – review & editing

Matthew T. Murrill: Data curation, Methodology, Formal analysis, Visualization, Writing – original draft

Katharine M. Tatum: Data curation, Project administration

Amina Ahmed: Data curation, Project administration, Writing – review & editing

Kaylynn Aiona: Data curation, Project administration, Writing – review & editing

Jeannette L. Aldous: Data curation, Project administration

Jennifer M. Flood: Conceptualization, Methodology, Funding acquisition

Michelle K. Haas: Data curation, Project administration, Writing – review & editing

Masahiro Narita: Data curation, Project administration, Writing – review & editing

April C. Pettit: Data curation, Project administration, Writing – review & editing

Marie N. Séraphin: Data curation, Project administration, Writing – review & editing

Adithya Cattamanchi: Conceptualization, Methodology, Writing – review & editing

Tracy L. Ayers: Conceptualization, Methodology

## Participating sites

Arizona Medical Clinic

Carolina Primary Care

Chinatown Public Health Center

Federico F. Pena Southwest Family Health Center

Grady Brookhaven Health Center

International Community Health Services

Kokua Kalihi Valley Comprehensive Family Services

North Park Family Medicine

San Ysidro Health Center

Siloam Health

South Florida Family Health and Research Center

St. Clare Medical Outreach

## Data availability statement

All individual participant data collected during the study, after deidentification, and additional documentation (data dictionary, data use guide and data extraction tools), are available at https://data.cdc.gov/National-Center-for-HIV-Viral-Hepatitis-STD-and-TB/Tuberculosis-Epidemiologic-Studies-Consortium-TBES/8xdf-byx4/about_data. Data are available indefinitely to anyone who wishes to access the data for any purpose.

---

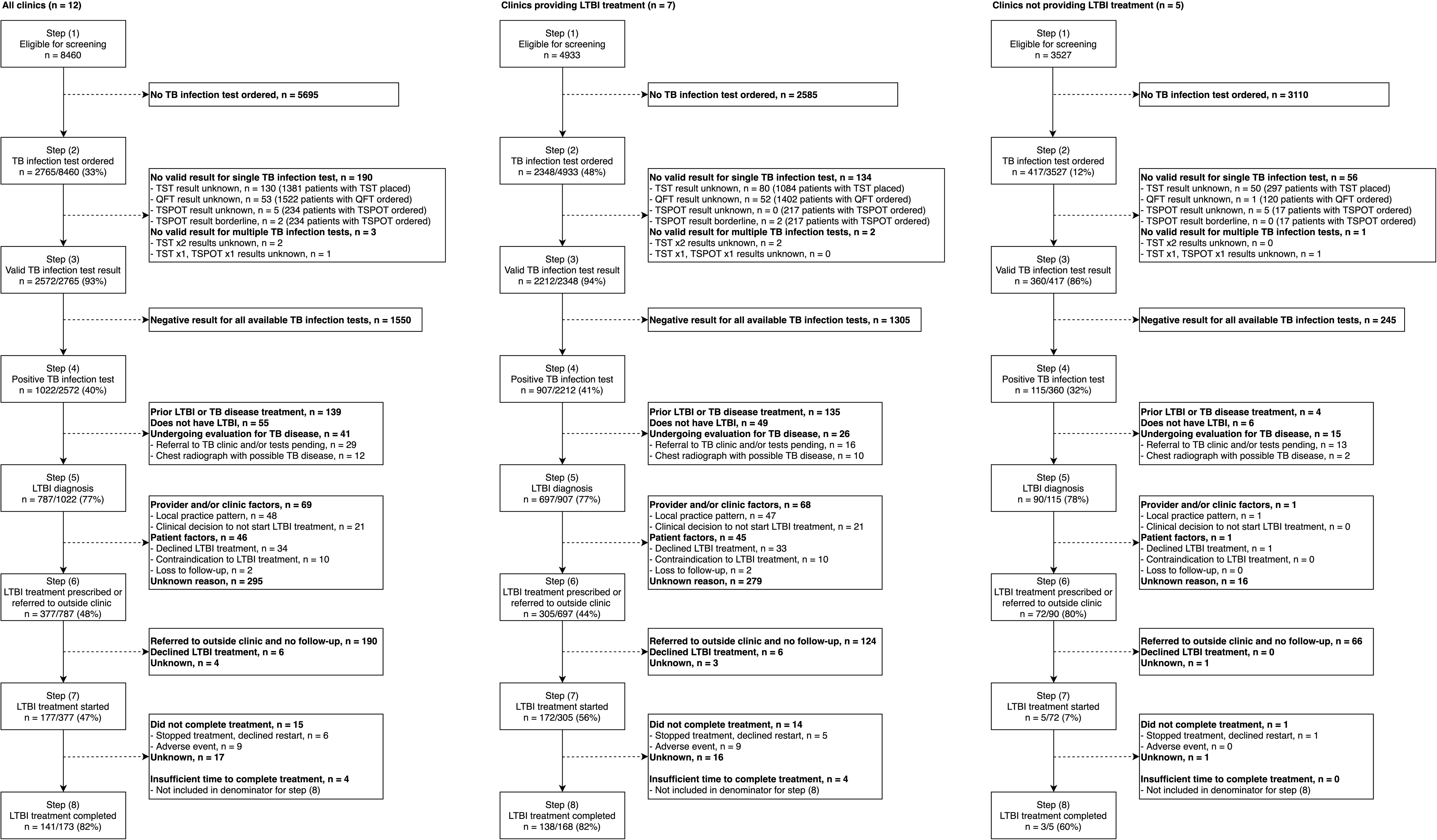

