## Supplementary Materials for "Identifying gaps in tuberculosis preventive care for non-U.S.-born persons at community health clinics in the United States"


**Authors and affiliations:**

Priya B. Shete*[1,2], Matthew T. Murrill[2,3]*, Katharine M. Tatum[4], Amina Ahmed[5,6], Kaylynn Aiona[7], Jeannette L. Aldous[8], Jennifer Flood[9], Michelle K. Haas[10,11], Masahiro Narita[12,13], April C. Pettit[14], Marie N. Séraphin[15,16], Adithya Cattamanchi[2,17], Tracy L. Ayers[18,19] for the Tuberculosis Epidemiologic Studies Consortium

*Authors contributed equally to this manuscript.

[1]Division of Pulmonary, Critical Care, Allergy and Sleep Medicine, University of California San Francisco, San Francisco, CA, USA

[2]Center for Tuberculosis, University of California San Francisco, San Francisco, CA, USA

[3]Division of Hospital Medicine, University of California San Francisco, San Francisco, CA, USA

[4]Northrop Grumman, Atlanta, GA, USA

[5]Levine Children's Hospital, Charlotte, North Carolina

[6]Department of Pediatrics, Wake Forest University School of Medicine, Winston-Salem, NC, USA

[7]Public Health Institute at Denver Health, Denver, CO, USA

[8]San Ysidro Health, San Diego, CA, USA

[9]Tuberculosis Control Branch, California Department of Public Health, Richmond, CA, USA

[10]Division of Mycobacterial and Respiratory Infections, Department of Medicine, National Jewish Health, Denver, CO, USA

[11]Division of Infectious Diseases, Department of Medicine, University of Colorado Anschutz Medical Campus, Aurora, CO, USA

[12]Public Heath Seattle & King County, Seattle, WA, USA

[13]Division of Pulmonary, Critical Care, and Sleep Medicine, University of Washington, Seattle, WA, USA

[14]Divisions of Infectious Diseases & Epidemiology, Department of Medicine, Vanderbilt University Medical Center, Nashville, TN, USA

[15]Division of Infectious Diseases and Global Medicine, Department of Medicine, University of Florida, Gainesville, FL, USA

[16]Emerging Pathogens Institute, University of Florida, Gainesville, FL, USA

[17]Division of Pulmonary Diseases and Critical Care Medicine, University of California Irvine, Irvine, USA

[18]Division of Tuberculosis Elimination, National Center for HIV, Viral Hepatitis, STD, and TB Prevention, Centers for Disease Control and Prevention, Atlanta, GA, USA

[19]Deceased author

*Cascade eligibility (cascade step 1)*

All non-U.S.-born individuals receiving routine care at participating community health clinics (CHCs) who met study eligibility criteria were included in this analysis. Inclusion criteria for this study were: 1) adult (≥18 years of age), 2) presentation for routine care on days of data collection, 3) established care at the participating clinic for ≥12 months and 4) non-U.S.-born, defined as birth outside of the US or a non-English primary language or use of an interpreter during a medical visit if place of birth was not available. Other risk factors for latent tuberculosis (TB) infection (LTBI), such as medical comorbidities, immunosuppression, residence in a congregate setting and persons experiencing homelessness, were not considered. Based on chart review, documentation of prior LTBI or TB disease diagnosis or treatment was not routinely available until after a positive TB infection test resulted and providers were considering LTBI treatment. To accurately capture observed clinical practice, individuals with prior LTBI or TB history were not excluded from the cascade at this step and still considered eligible for testing.

*TB infection testing (cascade steps 2-4)*

Three tests were routinely utilized by CHCs in the study: tuberculin skin test (TST) and two interferon-gamma release assays (IGRA) – QuantiFERON (QFT) and T-SPOT.*TB* (T-SPOT). Testing in the care cascade was divided into three steps. First, the ordering of a test was defined as the documentation of either the placement of tuberculin for TST or the ordering of a QFT or T-SPOT (Step 2). All relevant dates for testing were not uniformly available on chart review. Only qualitative test results were extracted (i.e. positive, negative, indeterminate, unknown). A TB infection test with a valid result was defined as a positive or negative qualitative result, excluding tests that were indeterminate or unknown (Step 3). Individuals were considered to have a positive test (Step 4) if they had ≥1 positive TB infection result of any type regardless of other test results.

*LTBI evaluation and diagnosis (cascade step 5)*

LTBI diagnosis was defined as having a positive TB infection test and no clinical documentation of prior LTBI treatment or TB disease diagnosis or treatment, no documentation of an outside clinic referral for TB evaluation, and no evidence of possible TB disease on available chest imaging identified during chart review. Due to the variable completion or reporting of chest imaging for TB disease evaluation, a normal chest radiograph was not required for LTBI diagnosis, but a chest radiograph suspicious for TB disease was used to exclude individuals that may have TB disease. Chest imaging studies were not reviewed by the study team, and the radiologist interpretation from routine clinical care was used. Data on TB symptom screening as a part of the medical evaluation for individuals with presumed LTBI was not available.

*LTBI treatment (cascade steps 6-8)*

Only a subset of participating CHCs provided both LTBI diagnostic and treatment services (7/12 clinics, 4933/8460 individuals). While documentation was identified for either the prescription of LTBI treatment or outside referral for LTBI treatment at all clinics (Step 6), documented LTBI treatment and follow-up (Step 7) was primarily only available for care provided at CHCs with LTBI treatment services. Treatment outcomes were determined by the chart review of clinical notes and not based on medication refills (Step 8).

*Statistical Analysis*

Individual-level covariates included age, sex, duration of care at clinic, whether an IGRA was used (only IGRA or in combination with TST), number of TB infection tests completed and LTBI treatment regimen. Clinic-level covariates included percent of clinic population without health insurance, any IGRA use at clinic, and clinic LTBI services available (testing only vs. treatment also prescribed by clinic providers). Due to large amounts of missingness that were highly variable by clinic, the evaluation of additional demographic data (place of birth, race and ethnicity) was not possible. Complete case analysis was performed for the model of TB infection test ordering with 9 individuals excluded due to missing age, sex or duration of care information.

Model diagnostics included examining both individual and clinic-level residual plots, sensitivity of analyses to the number of quadrature points used for model estimation as well as robust variance estimators. Due to the small number of individuals on LTBI treatment with known follow-up, sufficient time for treatment completion and seeking care at the 7 participating clinics where LTBI treatment was prescribed, only a limited set of factors could be evaluated in a model for treatment completion.

**Table E1:** Latent tuberculosis infection services currently provided by community health clinics

| Community health clinic | Individuals (n) | TST used | IGRA used | CXR on-site | LTBI Rx prescribed | Pharmacy on-site* |
|---|---|---|---|---|---|---|
| *LTBI diagnostic and treatment services* | | | | | | |
| Clinic 1 | 703 | Y | N | N | Y | N |
| Clinic 3 | 700 | Y | Y | N | Y | N |
| Clinic 4 | 714 | Y | Y | Y | Y | Y |
| Clinic 6 | 703 | Y | Y | N | Y | N |
| Clinic 7 | 709 | Y | N | N | Y | Y |
| Clinic 9 | 708 | Y | Y | Y | Y | Y |
| Clinic 12 | 700 | N | Y | N | Y | Y |
| *LTBI diagnostic services only* | | | | | | |
| Clinic 2 | 700 | Y | Y | Y | N | - |
| Clinic 5 | 735 | Y | N | Y | N | - |
| Clinic 8 | 700 | Y | Y | Y | N | - |
| Clinic 10 | 696 | Y | Y | Y | N | - |
| Clinic 11 | 696 | Y | Y | N | N | - |

Abbreviations:
- TB infection testing: TST = tuberculin skin testing, IGRA = interferon-gamma release assay
- CXR on-site = chest radiograph performed on-site
- LTBI Rx prescribed = LTBI treatment prescribed by clinic providers
- Y = service provided, N = service not provided

*Note: only clinics where providers prescribe LTBI treatment were asked if administration of treatment happens on site or if there was a pharmacy on site that can fill prescriptions.

**Table E2:** Tuberculosis infection tests currently used by clinic site (**Supplementary Materials: Clinic Assessment Form**) as well as documented TB infection tests in charts of individuals included in study (**Supplementary Materials: Patient Data Extraction Form** and review of entire clinical charts of selected individuals)

| Community health clinic | Individuals (n) | TST used | IGRA used | TST placed n (row %) | QFT ordered n (row %) | T-SPOT ordered n (row %) |
|---|---|---|---|---|---|---|
| Clinic 1 | 703 | Y | N | 104 (14.8%) | 99 (14.1%) | 65 (9.3%) |
| Clinic 2 | 700 | Y | Y | 53 (7.6%) | 43 (6.1%) | 2 (0.3%) |
| Clinic 3 | 700 | Y | Y | 108 (15.4%) | 634 (90.6%) | 0 |
| Clinic 4 | 714 | Y | Y | 23 (3.2%) | 248 (34.8%) | 0 |
| Clinic 5 | 735 | Y | N | 72 (9.8%) | 7 (1.0%) | 0 |
| Clinic 6 | 703 | Y | Y | 50 (7.1%) | 6 (0.9%) | 92 (13.1%) |
| Clinic 7 | 709 | Y | N | 414 (58.4%) | 2 (0.3%) | 0 |
| Clinic 8 | 700 | Y | Y | 77 (11.0%) | 21 (3.0%) | 0 |
| Clinic 9 | 708 | Y | Y | 99 (14.0%) | 171 (24.2%) | 0 |
| Clinic 10 | 696 | Y | Y | 50 (7.2%) | 45 (6.5%) | 0 |
| Clinic 11 | 696 | Y | Y | 45 (6.5%) | 4 (0.6%) | 15 (2.2%) |
| Clinic 12 | 700 | N | Y | 286 (40.9%) | 242 (34.6%) | 60 (8.6%) |
| **Total** | 8460 | - | - | 1381 (16.3%) | 1522 (18.0%) | 234 (2.8%) |

- Abbreviations: n = number of individuals, TST = tuberculin skin test, IGRA = interferon-gamma release assay, QFT = QuantiFERON, T-SPOT = T-SPOT.TB IGRA test, Y = service provided, N = service not provided

*Note: out of 2765 individuals who had at least 1 LTBI TST placed or IGRA ordered, 447 (16% of 2765) had >1 TST placed and/or IGRA ordered documented in their clinical chart. As a result, sum of the row percentages in columns TST placed, QFT ordered and TSPOT ordered can be more than 100%

**Table E3:** Individual and clinic-level factors associated with positive test result among individuals with at least one valid TB infection test result for non-U.S.-born individuals at 12 community health clinics in the US

| | **Summary statistics** | | | **Multilevel mixed logistic models** | | | | | |
|---|---|---|---|---|---|---|---|---|---|
| | n (column %) | n (row %) | n (row %) | Individual-level covariates | | | Individual and clinic-level covariates | | |
| **Individual-level variables** | All (n = 2572) | No positive TB infection test (n = 1550) | Positive TB infection test (n = 1022) | aOR | 95% CI | p | aOR | 95% CI | p |
| Age (years) | | | | | | | | | |
| ≤30 years | 301 (11.7%) | 239 (79.4%) | 62 (20.6%) | Ref | - | - | Ref | - | - |
| >30-50 years | 919 (35.7%) | 594 (64.6%) | 325 (35.4%) | 2.12 | [1.52,2.97] | <0.01 | 2.14 | [1.53,2.99] | <0.01 |
| >50-70 years | 1129 (43.9%) | 615 (54.5%) | 514 (45.5%) | 3.27 | [2.33,4.59] | <0.01 | 3.29 | [2.34,4.62] | <0.01 |
| >70 years | 223 (8.7%) | 102 (45.7%) | 121 (54.3%) | 4.41 | [2.87,6.78] | <0.01 | 4.46 | [2.90,6.85] | <0.01 |
| Sex | | | | | | | | | |
| Male | 848 (33.0%) | 465 (54.8%) | 383 (45.2%) | Ref | - | - | Ref | - | - |
| Female | 1724 (67.0%) | 1085 (62.9%) | 639 (37.1%) | 0.74 | [0.62,0.89] | 0.001 | 0.74 | [0.62,0.89] | 0.001 |
| Duration of care at clinic (years), median (IQR) | 5.25 (3.01, 7.86) | 4.81 (2.70, 7.28) | 6.25 (3.73, 8.67) | 1.06 | [1.03,1.09] | <0.01 | 1.06 | [1.03,1.09] | <0.01 |
| Number of TB infection tests ordered | | | | | | | | | |
| 1 test | 2128 (82.7%) | 1392 (65.4%) | 736 (34.6%) | Ref | - | - | Ref | - | - |
| ≥2 tests | 444 (17.3%) | 158 (35.6%) | 286 (64.4%) | 4.27 | [3.36,5.43] | <0.01 | 4.25 | [3.34,5.40] | <0.01 |
| **Clinic-level variables** | | | | | | | | | |
| Clinic population without insurance | | | | | | | | | |
| <25% (n = 8) | 1828 (71.1%) | 1122 (61.4%) | 706 (38.6%) | | | | Ref | - | - |
| ≥25% (n = 4) | 744 (28.9%) | 428 (57.5%) | 316 (42.5%) | | | | 1.56 | [0.81,2.99] | 0.18 |

Abbreviations: n = number of individuals, aOR = adjusted odds ratio, 95% CI = 95% confidence interval, p = p-value, IQR = interquartile range, IGRA = interferon gamma release assay

**Table E4:** Chest radiograph currently available on-site (**Supplementary Materials: Clinic Assessment Form**) as well as availability in the medical record of a chest radiograph used to evaluate for tuberculosis disease after a positive TB infection test result (**Supplementary Materials: Patient Data Extraction Form** )

| Community health clinic | Individuals with positive TB infection test | Chest radiograph available on-site | Chest radiograph result available* | Chest radiograph result*: Normal | Chest radiograph result*: Abnormal (not suspicious for TB) | Chest radiograph result*: Abnormal (suspicious for TB) | No chest radiograph result available* |
|---|---|---|---|---|---|---|---|
| Clinic 1 | 94 | N | 49 (52.1%) | 31 (33.0%) | 15 (16.0%) | 3 (3.2%) | 45 (47.9%) |
| Clinic 2 | 28 | Y | 23 (82.1%) | 22 (78.6%) | 1 (3.6%) | 0 (0.0%) | 5 (17.9%) |
| Clinic 3 | 275 | N | 176 (64.0%) | 157 (57.1%) | 15 (5.5%) | 4 (1.5%) | 99 (36.0%) |
| Clinic 4 | 60 | Y | 52 (86.7%) | 43 (71.7%) | 6 (10.0%) | 3 (5.0%) | 8 (13.3%) |
| Clinic 5 | 34 | Y | 18 (52.9%) | 17 (50.0%) | 0 (0.0%) | 1 (2.9%) | 16 (47.1%) |
| Clinic 6 | 69 | N | 59 (85.5%) | 45 (65.2%) | 11 (15.9%) | 3 (4.4%) | 10 (14.5%) |
| Clinic 7 | 160 | N | 133 (83.1%) | 125 (78.1%) | 6 (3.8%) | 2 (1.3%) | 27 (16.9%) |
| Clinic 8 | 19 | Y | 9 (47.4%) | 8 (42.1%) | 0 (0.0%) | 1 (5.3%) | 10 (52.6%) |
| Clinic 9 | 108 | Y | 78 (72.2%) | 71 (65.7%) | 5 (4.6%) | 2 (1.9%) | 30 (27.8%) |
| Clinic 10 | 33 | Y | 15 (45.5%) | 13 (39.4%) | 2 (6.1%) | 0 (0.0%) | 18 (54.6%) |
| Clinic 11 | 1 | N | 1 (100.0%) | 1 (100.0%) | 0 (0.0%) | 0 (0.0%) | 0 (0.0%) |
| Clinic 12 | 141 | N | 92 (65.2%) | 86 (61.0%) | 5 (3.6%) | 1 (0.7%) | 49 (34.8%) |
| **Total** | 1022 | - | 705 (69.0%) | 619 (60.6%) | 66 (6.5%) | 20 (2.0%) | 317 (31.0%) |

- Abbreviations: TB = tuberculosis, Y = service provided, N = service not provided
*Row % of individuals with positive TB infection test

**Table E5:** Latent tuberculosis infection care cascade steps by patterns of test results for individuals with two or more tests with valid results

| Patterns of latent tuberculosis test results | | Latent tuberculosis infection care cascade steps* | | | | |
|---|---|---|---|---|---|---|
| | | Step 5: LTBI diagnosis | Step 6: Treatment offered or referred to outside clinic | Step 7a: Treatment offered and follow-up information available | Step 7b: Treatment started | Step 8: Treatment completed |
| **2 tests**† | TST +/+ | 7 | 2 (29%) | 1 (50%) | 1 (100%) | 1 (100%) |
| | TST +/- | 6 | 2 (33%) | 1 (50%) | 1 (100%) | 0 (0%) |
| | IGRA +/+ | 5 | 2 (40%) | 1 (50%) | 1 (100%) | 1 (100%) |
| | IGRA +/- | 1 | 1 (100%) | 0 (0%) | | |
| | TST+/IGRA+ | 93 | 70 (75%) | 24 (34%) | 23 (96%) | 16 (70%) |
| | TST+/IGRA- | 46 | 2 (4%) | 1 (50%) | 1 (100%) | 0 (0%) |
| | TST-/IGRA+ | 5 | 3 (60%) | 1 (33%) | 1 (100%) | 1 (100%) |
| **3+ tests**‡ | TST+/IGRA+ (≥3 tests) | 16 | 11 (69%) | 1 (9%) | 1 (100%) | 1 (100%) |
| | TST+/IGRA- x2 | 8 | 2 (25%) | 0 (0%) | | |
| | TST-/IGRA+ x2 | 2 | 2 (100%) | 0 (0%) | | |
| | Other patterns | 5 | 3 (60%) | 1 (33%) | 1 (100%) | 1 (100%) |
| **All individuals with ≥2 valid tests** | | 194 | 100 (52%) | 31 (31%) | 30 (97%) | 21 (70%) |

Abbreviations:
- TST = tuberculin skin test
- IGRA = interferon gamma release assay test (i.e. QuantiFERON or T-SPOT.TB tests)

*Data provided for each cascade step and pattern of test results:
### of individuals completing each step
(% of individuals in previous step completing current step)

†Testing patterns: 2 tests with valid results
+/+ = both tests positive
+/- = one test positive and one test negative
-/- = both tests negative

‡Testing patterns: 3+ tests with valid results
TST+/IGRA+ (≥3 tests) = three or more tests (TST and/or IGRA) all with positive results
TST+/IGRA- x2 = one positive TST and 2 negative IGRAs
TST-/IGRA+ x2 = one negative TST and 2 positive IGRAs

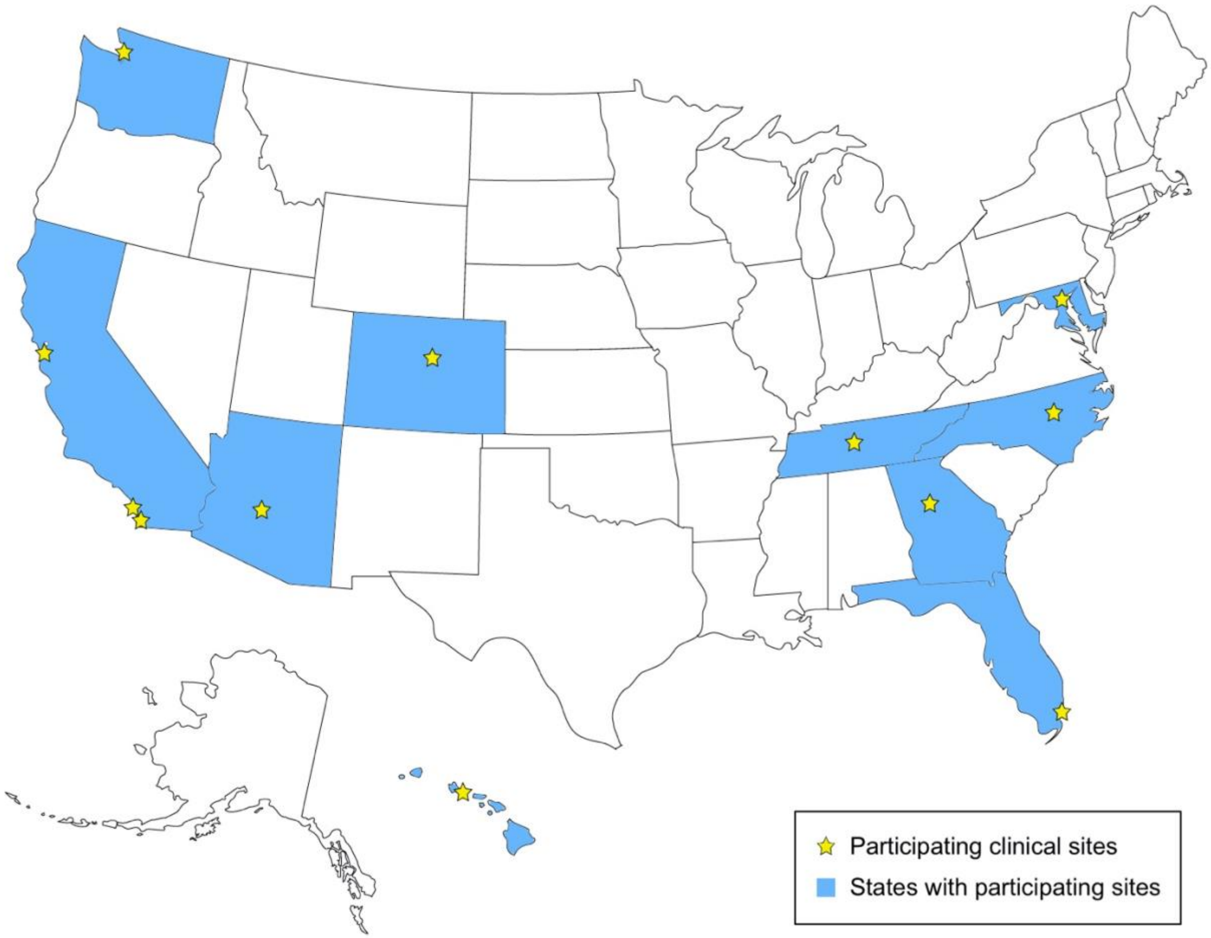


**Figure E1:** Community health clinics (CHCs) participating in Tuberculosis Epidemiologic Studies Consortium 2, Part C study (n = 12). Stars indicate locations of the participating CHCs, spanning 10 US states (highlighted in blue)

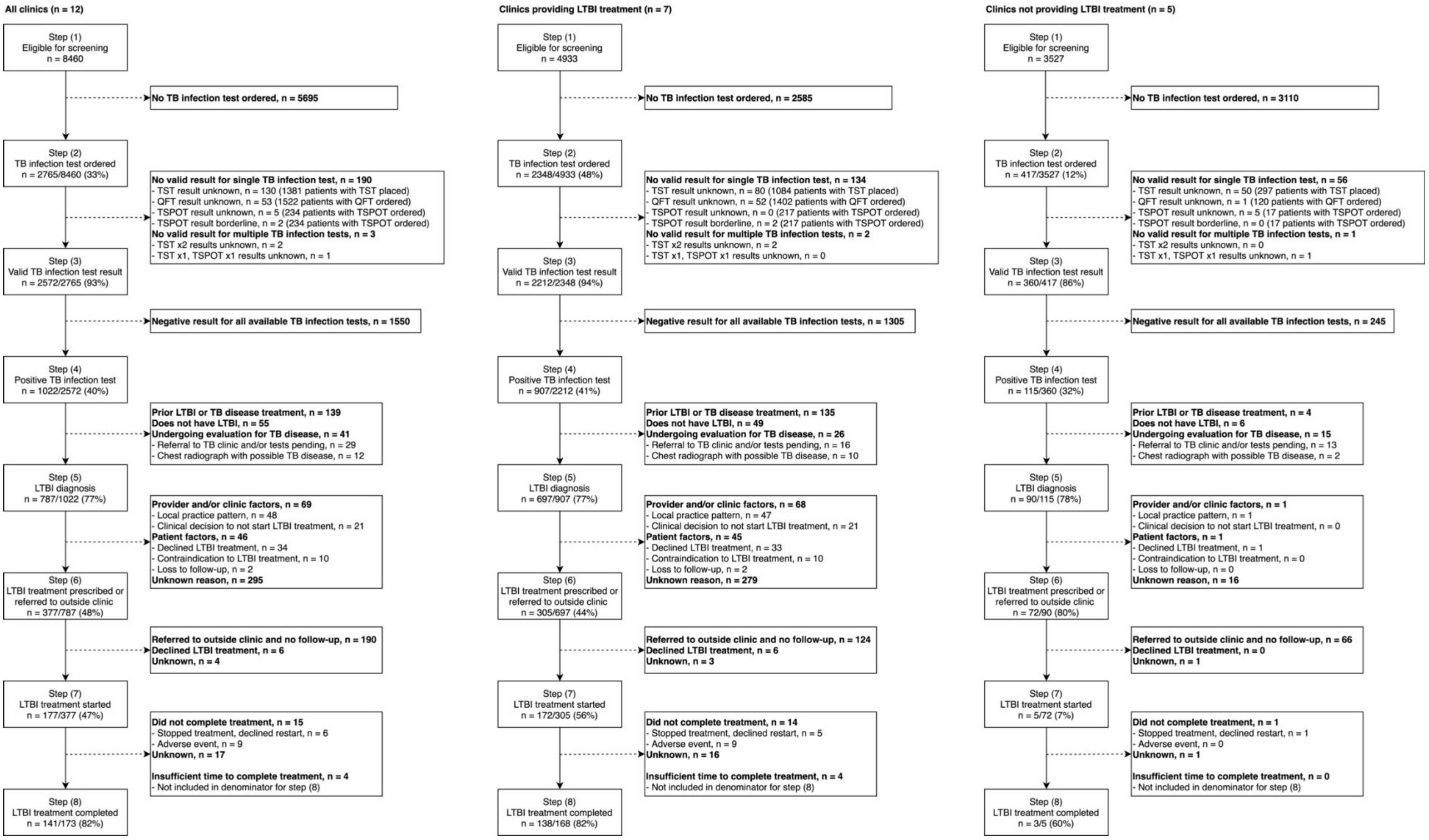


**Figure E2:** Eligibility and latent TB infection (LTBI) care cascade for non-U.S.-born individuals at 12 participating community health clinics across the US (**left panel**). Stratified eligibility and LTBI care cascades by LTBI clinic services also shown -- community health clinics providing LTBI diagnostic and treatment services (**middle panel**, n = 7) and community health clinics providing only LTBI diagnostic services (**right panel**, n = 5). Abbreviations: n = number of individuals, TB = tuberculosis, LTBI = latent tuberculosis infection, tuberculin skin testing (TST), QuantiFERON (QFT), T-SPOT (T-SPOT.TB test).

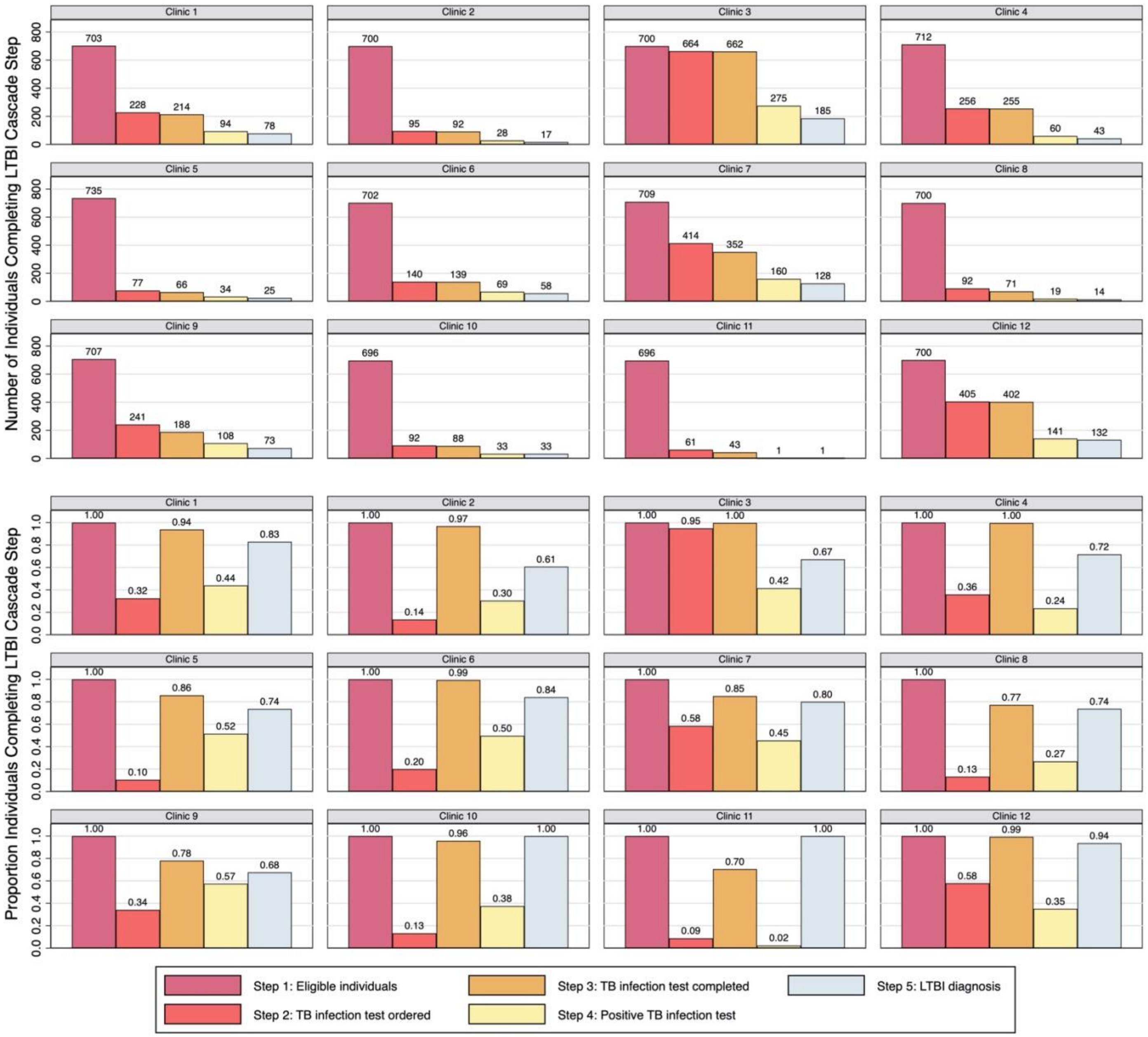


**Figure E3:** Latent TB infection (LTBI) diagnostic care cascade stratified by community health clinic (CHC), number of individuals completing each step (top panel) and proportion of individuals completing sequential steps (bottom panel)

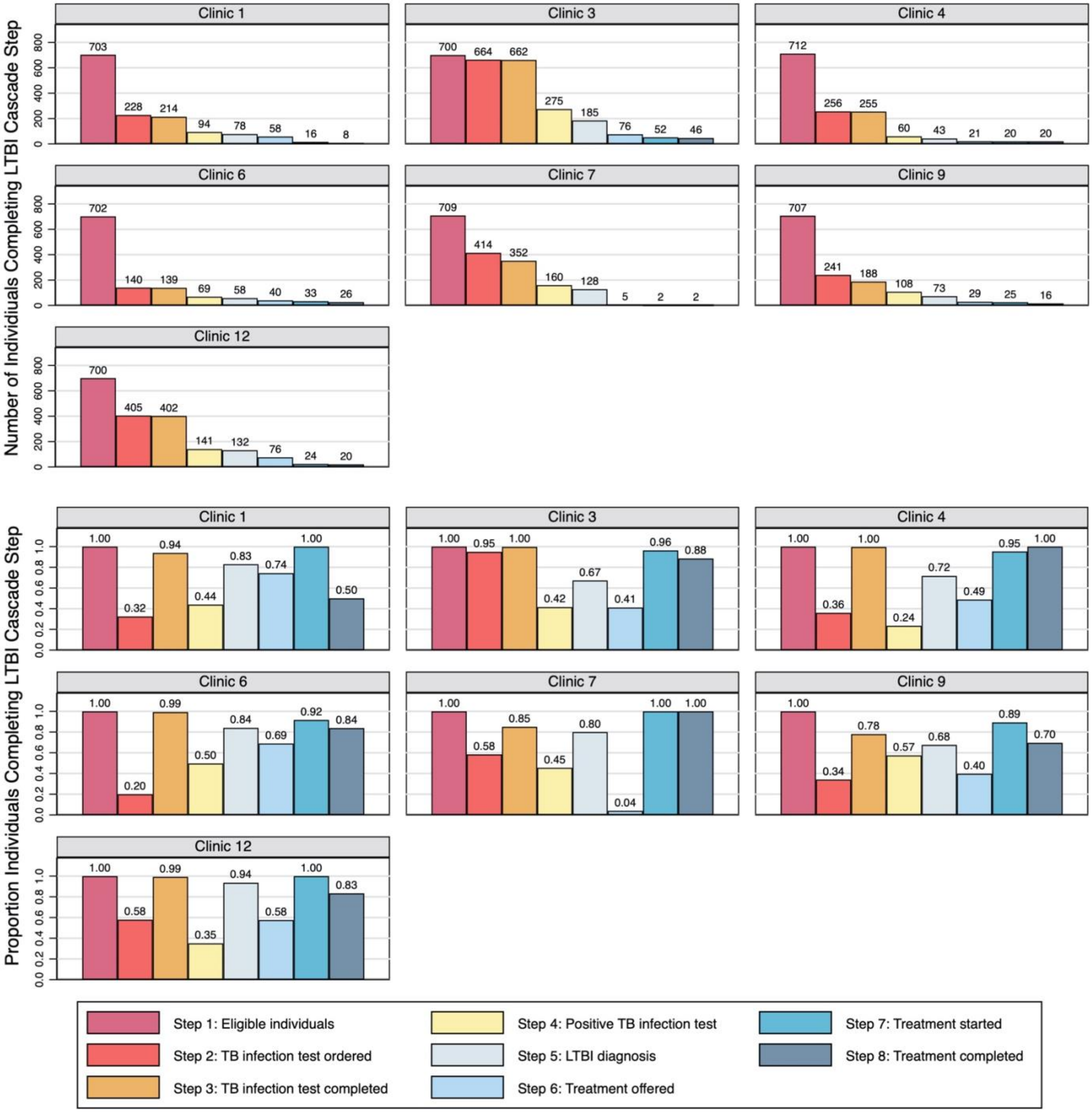


**Figure E4:** Latent TB infection (LTBI) diagnostic and treatment care cascade stratified by community health clinic (CHC) for CHCs that offer LTBI treatment services. Number of individuals completing each step (top panel) and proportion of individuals completing sequential steps (bottom panel)

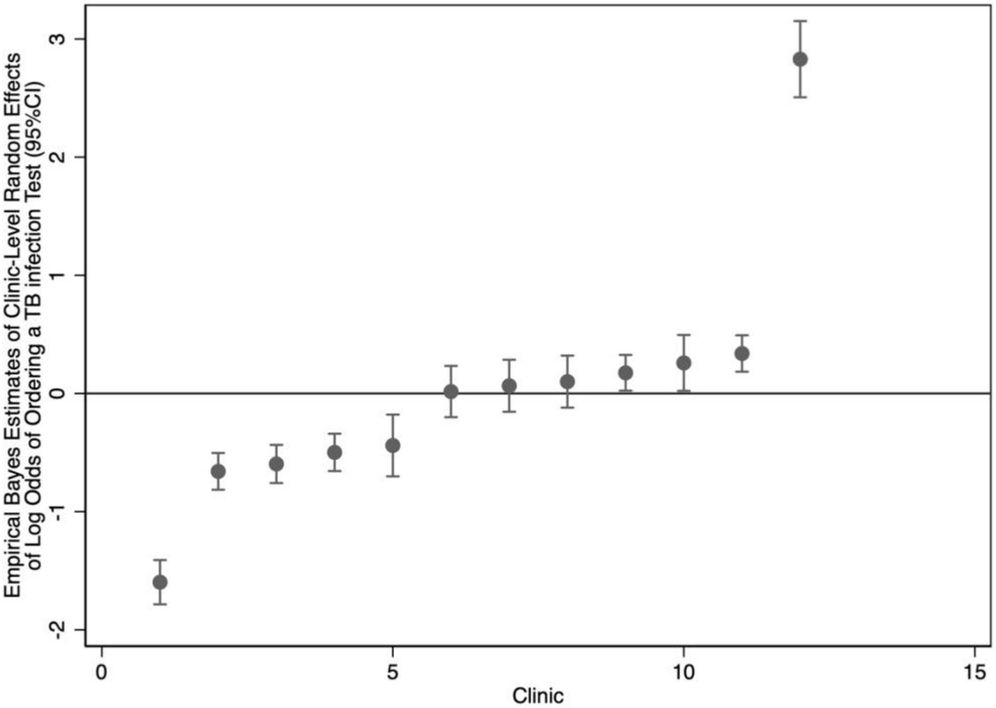


**Figure E5:** Empirical Bayes estimates of the clinic-level deviation of the average log odds of ordering a TB infection test (among those eligible for testing) from the overall average for the study population

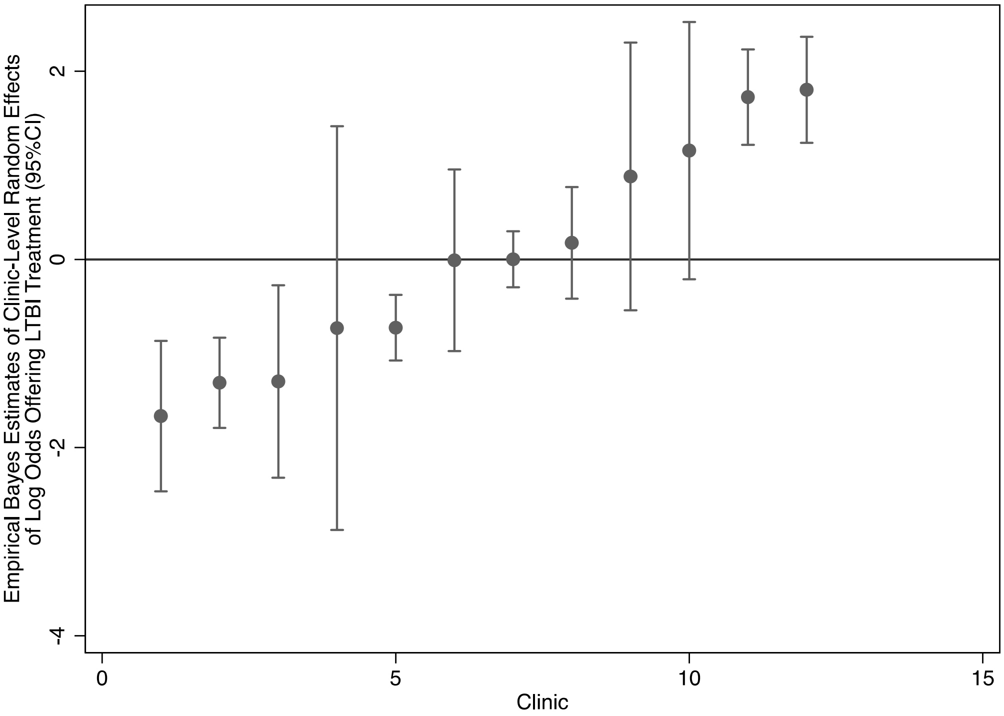


**Figure E6:** Empirical Bayes estimates of the clinic-level deviation of the average log odds of offering latent tuberculosis infection (LTBI) treatment (among those with LTBI diagnosis) from the overall average for the study population

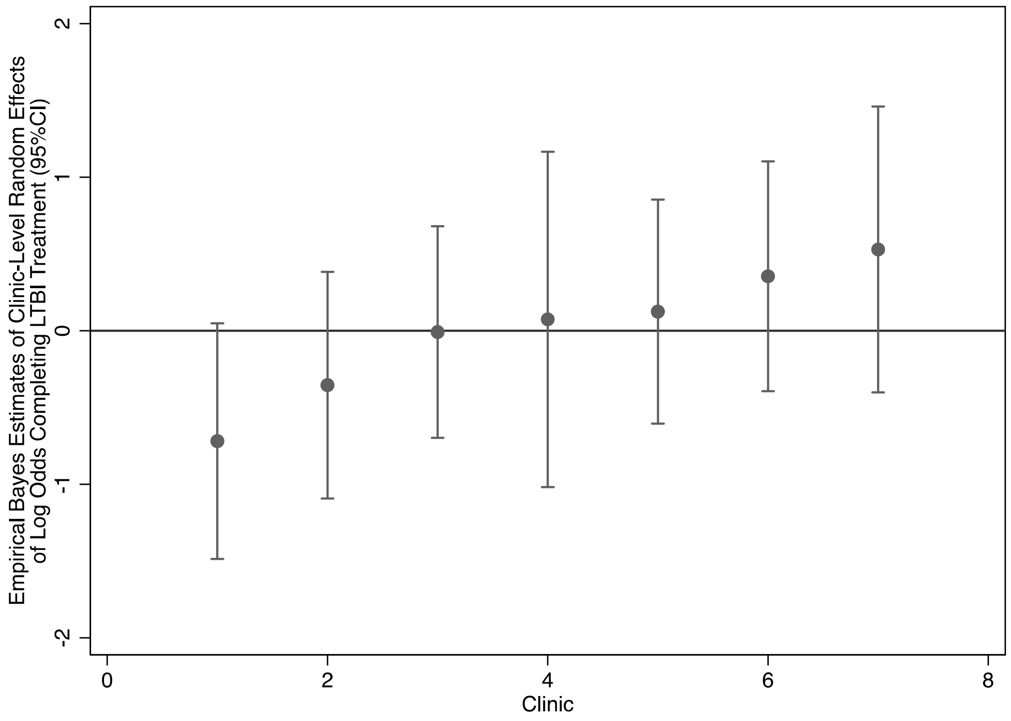


**Figure E7:** Empirical Bayes estimates of the clinic-level deviation of the average log odds of completing latent tuberculosis infection (LTBI) treatment (among those who started treatment and had sufficient time for completion) from the overall average for the study population
