## Supplementary material for "Identifying gaps in tuberculosis preventive care for non-U.S.-born persons at community health clinics in the United States": Clinic Assessment Form

### Appendix B: Latent Tuberculosis Infection (LTBI) Detailed Clinic Assessment

This questionnaire will help us understand your clinic's current approach to LTBI testing and treatment. We will use this information to inform our approach to partnerships with community clinics like yours. All of the information you provide will be confidential and will not be shared outside of TBESC; it will only be used for research purposes.

1. TBESC site: ___________________

2. Clinic name: ___________________

---

#### Clinic characteristics

3. Which specialties are practiced at your clinic? *[Check all that apply]*
   ☐ Family medicine
   ☐ Internal medicine
   ☐ Obstetrics and gynecology
   ☐ Pediatrics
   ☐ Other, specify:_________________________________________

4. Approximately what proportion of your patients do NOT have any form of health insurance *(i.e., are completely self-pay or no pay)*?
   ☐ <25%
   ☐ 25-50%
   ☐ >50%
   ☐ Unknown

5. What services does your clinic offer for free if your patients are unable to pay? *[Check all that apply]*
   ☐ Clinic visits
   ☐ Tuberculin skin test (TST)
   ☐ T-SPOT.*TB* test
   ☐ QuantiFERON-TB (QFT) test
   ☐ Chest X-ray (CXR)
   ☐ Chest CT scan
   ☐ LTBI treatment medications
   ☐ Blood-based laboratory tests

---

#### LTBI testing practices

6. Does your clinic use the tuberculin skin test (TST) for LTBI?
   ☐ Yes
   ☐ No

7. Does your clinic use the QuantiFERON-TB (QFT) test for LTBI? *(NOTE: this refers to both QFT Gold In-tube and QFT-Plus)*
   ☐ Yes
   ☐ No *(Skip to Question 8)*

   a. Does your clinic perform the <u>blood draw</u> on site?
      ☐ Yes
      ☐ No

   b. Which laboratory performs the test?
      ☐ On-site laboratory
      ☐ Quest Diagnostics
      ☐ LabCorp
      ☐ Other laboratory, specify:______________________________
      ☐ Unknown

8. Does your clinic use the T-SPOT.*TB* (TSPOT) test for LTBI?
   ☐ Yes
   ☐ No *(Skip to Question 9)*

   a. Does your clinic perform the <u>blood draw</u> on site?
      ☐ Yes
      ☐ No

   b. Which laboratory performs the test?
      ☐ On-site laboratory
      ☐ Oxford Immunotec
      ☐ Other laboratory, specify:______________________________
      ☐ Unknown

9. Are chest radiographs performed on site?
   ☐ Yes
   ☐ No

   a. Are the chest radiograph reports made available to the clinic electronically?
      ☐ Yes
      ☐ No

---

#### LTBI treatment practices

10. Do providers at your clinic prescribe LTBI treatment?
    ☐ Yes
    ☐ No *(Skip to Question 12)*

    a. Which LTBI treatment regimens are prescribed by clinicians at your clinic? *(Check all that apply)*
       ☐ Isoniazid, 9 months
       ☐ Isoniazid, 6 months

☐ Isoniazid and rifapentine, 12 weekly doses
☐ Rifampin, 4 months
☐ Other, specify: ___________________________________

11. Does your clinic administer treatment on site (is there an onsite pharmacy that can fill prescriptions)?
☐ Yes *(Skip to Question 14)*
☐ No

12. Where does your clinic refer patients for LTBI treatment? *(Check all that apply)*
☐ Health department
☐ Federally qualified health center (FQHC)
☐ Private clinic
☐ Other, specify:___________________________________
☐ Patients are not referred elsewhere for LTBI treatment
☐ Unknown

13. What are the barriers to providing treatment at your clinic? *(Check all that apply)*
☐ Not enough staff available to oversee treatment completion
☐ Not enough money to purchase drugs
☐ No pharmacist on staff
☐ Insurance does not cover LTBI treatment
☐ Not reimbursed well enough
☐ Tuberculosis is not a pertinent problem for our clinic
☐ Other, specify:___________________________________
☐ Unknown

---

#### Medical records

14. What kind of medical record system does your clinic currently use? *(Check all that apply)*
☐ Paper
☐ Epic
☐ Cerner
☐ NextGen
☐ eClinicalWorks
☐ Other electronic medical record system, specify:___________________________________

15. During a typical clinic visit, if a clinician wanted to search a patient's electronic medical record how many years back could they search?
☐ < 1 year
☐ 1-3 years
☐ 4-5 years
☐ 5-10 years
☐ > 10 years

16. What information is recorded in your medical records? *[Check all that apply}*

☐ Country of birth
☐ Language preference
Language information collected:________________________________
☐ Interpreter needed
Interpreter information collected: ________________________________
☐ Tuberculosis (TB) signs or symptoms
☐ TST placement date
☐ TST read date
☐ TST result
☐ QFT blood draw date
☐ QFT result
☐ TSPOT blood draw date
☐ TSPOT result
☐ Date chest radiograph performed
☐ Chest radiograph results
☐ Date chest CT scan performed
☐ Chest CT scan results
☐ History of TB or LTBI treatment
☐ LTBI treatment regimen
☐ LTBI treatment start date
☐ LTBI treatment stop date
☐ Reason LTBI treatment not completed

17. Is country of birth routinely collected for all patients?
☐ Yes
☐ No

---

#### Written policies

18. Does your clinic have written policies on LTBI screening, testing, and/or treatment?
☐ Yes *(Upload document)*
☐ No

---

#### Future collaboration

19. Please provide any additional information that would help us in assisting you to increase LTBI screening, testing and treatment at your clinic.
