## Supplementary material for "Identifying gaps in tuberculosis preventive care for non-U.S.-born persons at community health clinics in the United States": Patient Data Extraction Form

### APPENDIX C: PATIENT DATA EXTRACTION TOOL

*[Text written in italics between brackets is only meant for the tool creator as guidance. It will not be displayed to those entering data into REDCap]*

1. TBESC site: ________________

2. Clinic name: ________________

---

**Patient Demographic and Clinical Characteristics**

3. How was this patient determined to be eligible for purposes of this study? *(Check all that apply)*
   ☐ Born outside of the United States
   Select patient's country of birth: ________________________ *[drop-down list]*
   ☐ Primary language other than English
   Select patient's primary language: ________________________ *[drop-down list]*
   ☐ Patient required interpreter

4. Date of current/most recent visit: [MM/DD/YYYY]

5. Date of first visit: [MM/DD/YYYY]

6. Was the patient's date of birth or age collected?
   ☐ Yes
   Patient's current age (in years): [###]
   ☐ No

7. What is the patient's sex?
   ☐ Male
   ☐ Female
   ☐ Unknown

8. What is the patient's race? *(Check all that apply)*
   ☐ American Indian/Alaska Native
   ☐ Asian________________________ [*drop-down list*]
   ☐ Black/African American
   ☐ White/Caucasian
   ☐ Native Hawaiian/Pacific Islander
   ☐ Unknown

9. What is the patient's ethnicity?
   ☐ Hispanic or Latino
   ☐ Not Hispanic or Latino
   ☐ Unknown

*For questions 10-15, if information for the test is not recorded in the chart, select "No".*

#### LTBI Testing

10. Did this patient have a tuberculin skin test (TST) result from outside this clinic?
    ☐ Yes
    ☐ No *(Skip to Question 11)*

    *Complete the following questions for the most recent TST result from outside this clinic.*

    a. Is the date TST placed available?
       ☐ Yes
       Date TST placed: [MM/DD/YYYY]
       ☐ No

    b. Is the date TST read available?
       ☐ Yes
       Date TST read: [MM/DD/YYYY]
       ☐ No

    c. TST interpretation:
       ☐ Positive
       ☐ Negative
       ☐ Unknown

11. Did the patient have a QuantiFERON-TB (QFT) test ordered *(refers to both QFT Gold In-tube and QFT-Plus)* outside the clinic?
    ☐ Yes
    ☐ No *(Skip to Question 12)*

    *Complete the following questions for the most recent QFT test ordered outside this clinic.*

    a. Is the date of QFT blood draw available?
       ☐ Yes
       Date of QFT blood draw: [MM/DD/YYYY]
       ☐ No

    b. QFT interpretation:
       ☐ Positive
       ☐ Negative
       ☐ Indeterminate

☐ Failed
☐ Unknown

12. Did this patient have a T-SPOT.*TB* (TSPOT) test ordered outside this clinic?
☐ Yes
☐ No *(Skip to Question 13)*

*Complete the following questions for the most recent TSPOT test ordered outside this clinic.*

a. Is the date of TSPOT blood draw available?
☐ Yes
Date of TSPOT blood draw: [MM/DD/YYYY]
☐ No

b. TSPOT interpretation:
☐ Positive
☐ Negative
☐ Borderline
☐ Invalid
☐ Test not performed
☐ Unknown

13. Did this patient have a tuberculin skin test (TST) result at this clinic?
☐ Yes
☐ No *(Skip to Question 14)*

*Complete the following questions for the most recent TST result at this clinic.*

a. Is the date TST placed available?
☐ Yes
Date TST placed: [MM/DD/YYYY]
☐ No

b. Is the date TST read available?
☐ Yes
Date TST read: [MM/DD/YYYY]
☐ No

c. TST interpretation:
☐ Positive
☐ Negative
☐ Unknown

14. Did this patient have a QuantiFERON-TB (QFT) test *(refers to both QFT Gold In-tube and QFT-Plus)* ordered at this clinic? (regardless of what laboratory the sample was sent to)
☐ Yes
☐ No *(Skip to Question 15)*

*Complete the following questions for the most recent QFT test ordered at this clinic.*

a. Is the date of QFT blood draw available?
   ☐ Yes
   Date of QFT blood draw: [MM/DD/YYYY]
   ☐ No

b. QFT interpretation:
   ☐ Positive
   ☐ Negative
   ☐ Indeterminate
   ☐ Failed
   ☐ Unknown

15. Did this patient have a T-SPOT.*TB* (TSPOT) test ordered at this clinic? (regardless of what laboratory the sample was sent to)
    ☐ Yes
    ☐ No *(Skip to Question 16)*

*Complete the following questions for the most recent TSPOT test ordered at this clinic.*

a. Is the date of TSPOT blood draw available?
   ☐ Yes
   Date of TSPOT blood draw: [MM/DD/YYYY]
   ☐ No

b. TSPOT interpretation:
   ☐ Positive
   ☐ Negative
   ☐ Borderline
   ☐ Invalid
   ☐ Test not performed
   ☐ Unknown

16. Was a chest X-ray (CXR) done to rule out TB disease for this patient? [*Only displayed with evidence of a positive LTBI test above]*
    ☐ Yes
    ☐ No *(Skip to Question 17))*
    ☐ LTBI test result(s) pending *(STOP)*
    ☐ Unknown

*Complete the following questions for the most recent CXR following a positive LTBI above.*

a. Is the date of CXR available?
   ☐ Yes
   Date of CXR: [MM/DD/YYYY]
   ☐ No

b. CXR result:
   ☐ Normal
   ☐ Abnormal (not suspicious for TB)
   ☐ Abnormal (suspicious for TB)
   ☐ Unknown

**LTBI Treatment** *[Only asked if patient has a documented positive test above]*

17. Did a provider at the clinic prescribe LTBI treatment to the patient?
   ☐ Yes
   ☐ No *(Skip to Question 17b)*
   ☐ LTBI diagnosis pending *(STOP)*
   ☐ Patient sought/is seeking treatment elsewhere *(STOP)*

   *[Complete the following question if a provider prescribed LTBI treatment, then skip to Question 18]*

   a. Which LTBI treatment regimen was prescribed?
      ☐ Isoniazid, 9 months
      ☐ Isoniazid, 6 months
      ☐ Isoniazid and rifapentine, 12 weekly doses
      ☐ Rifampin, 4 months
      ☐ Other, specify:______________
      ☐ Unknown

   *[Complete the following question if a provider did not prescribe LTBI treatment, then STOP]*

   b. Why was LTBI treatment not prescribed?
      ☐ Patient refused treatment
      ☐ Patient was previously treated for LTBI/TB
      ☐ Patient had a contraindication
      ☐ Patient was lost to follow up
      ☐ Local practice
      ☐ Clinician decision
      ☐ Other, specify:____________
      ☐ Patient does not have LTBI
      ☐ Unknown
      *(STOP)*

18. Did the patient start LTBI treatment?
☐ Yes
☐ No (*Skip to Question 18d)*

*[Complete the following questions a-c if the patient started LTBI treatment, then skip to Question 19]*

a. Which LTBI treatment regimen was started?
☐ Isoniazid, 9 months
☐ Isoniazid, 6 months
☐ Isoniazid and rifapentine, 12 weekly doses
☐ Rifampin, 4 months
☐ Other, specify:________________
☐ Unknown

b. Are the number of prescriptions filled by this patient for the current treatment regimen available?
☐ Yes
Total number of prescriptions filled: [###]
☐ No *(Skip to Question 19)*

c. Are the number of doses per prescription for the current treatment regimen available?
☐ Yes
Total number of doses filled *(# of prescriptions x # of doses per prescription)*: [###]
☐ No
*(Skip to Question 19)*

*[Complete the following question if the patient did not start LTBI treatment, then STOP]*

d. Why did the patient not start LTBI treatment?
☐ Patient declined treatment
☐ Patient did not pick up prescription or medication
☐ Patient was lost to care
☐ Patient died
☐ Other, specify:_______________
☐ Unknown

19. Did the patient complete LTBI treatment?
☐ Yes *(STOP)*
☐ No
☐ Patient has not had sufficient time to complete LTBI treatment *(STOP)*
☐ Unknown *(STOP)*

*[Complete the following question if the patient did not complete LTBI treatment]*

a. Why did the patient not complete LTBI treatment?

☐ Patient declined further treatment
☐ Patient did not pick up prescription or medication
☐ Adverse event
☐ Patient became pregnant
☐ Patient died
☐ Patient moved
☐ Other, specify:________________
☐ Unknown
